# Genome-wide meta-analysis and integrative fine-mapping identify novel susceptibility loci and effector genes in psoriatic arthritis

**DOI:** 10.1101/2025.08.26.25334362

**Authors:** Yask Gupta, Tatiana Sezin, Diamant Thaçi

**Author notes:** Correspondence: Tatiana Sezin, Institute and Comprehensive Center for Inflammation Medicine, University of Lübeck, 23538, Lübeck, Germany. shared last author (Tatiana Sezin, Diamant Thaçi).

## Abstract

Psoriatic arthritis (PsA) is a complex immune-mediated inflammatory disorder affecting both the skin and musculoskeletal system. Although genome-wide association studies (GWAS) have identified multiple loci shared with psoriasis (PsO), the unique genetic architecture of PsA remains incompletely understood. To define PsA-specific genetic risk, we conducted a GWAS meta-analysis of 13,512 PsA cases and 715,639 non-PsA population controls, identifying 40 susceptibility loci, including 21 novel loci outside the MHC region. These include putative coding variants in genes such as *DENND1B*, as well as non-coding variants implicating new therapeutic targets. We applied expression quantitative trait loci (eQTL)-based post-GWAS analyses to prioritize high-confidence candidate genes at both novel (e.g., *PRR5L*) and established loci. To further resolve cellular context, we curated and re-analyzed a comprehensive, cell type–resolved eQTL resource, enabling downstream single-cell analyses that identified disease-relevant NADK-positive monocyte subsets and implicated candidate genes such as *ETS1*. Finally, we uncovered PsA-specific genetic signals within PsO GWAS loci, including a previously unrecognized association at *C1orf14* within the *IL23R* region. Our findings underscore the importance of large-scale genetic studies combined with fine phenotypic resolution to disentangle shared and disease-specific genetic mechanisms in PsA.

## Introduction

Psoriatic arthritis (PsA) is a chronic inflammatory disease affecting both joints and skin, often causing significant disability, reduced quality of life, and increased mortality^1^. It is characterized by musculoskeletal inflammation in different domains manifesting as arthritis, enthesitis, spondylitis, and dactylitis^2,3^. PsA typically arises in middle-aged individuals and confers elevated risk for cardiovascular disease, uveitis, metabolic syndrome, gastrointestinal disorders, and psychiatric comorbidities among others^1,4–7^. Affecting up to 0.25% of the general population, PsA develops in approximately 30% of patients with psoriasis (PsO)^1,2,8^. Although skin and nail involvement are present in up to 90% of PsA cases, PsA can also manifest independently, reflecting a complex, heterogeneous pathophysiology involving both innate and adaptive immune responses^2,9^.

PsA has a strong genetic component, with heritability estimates of 50–70% and a 30–55% recurrence risk among first-degree relatives^10–13^. Accordingly, genome-wide association studies (GWAS) have identified multiple risk loci, many overlapping with PsO but also PsA-specific signals implicating joint inflammation and immune dysregulation^14–18^. The major histocompatibility complex (MHC) class I region remains the strongest genetic risk factor in PsA, while non-MHC loci highlight key immune pathways including IL-23/IL-17, TNF-alpha, and NF-κB-consistent with the success of targeted therapies PsO and PsA^19–25^.

Despite recent advances in understanding the genetic architecture of PsA, the majority of GWAS-identified variants remain functionally uncharacterized, and their causal roles, especially in disease-relevant immune cells is still largely unknown^26^. Thus, a major challenge in the field lies in resolving the causal variants within associated loci and understanding how these variants influence gene regulation in PsA-specific immune contexts^27^.

To this end, we performed a large-scale GWAS meta-analysis in individuals of European ancestry. We identified novel risk loci and refined putative causal variant mapping in PsA. Furthermore, we integrated this data with a comprehensive single-cell blood eQTL dataset derived from multiple cohorts, enabling linkage of genetic risk to specific immune cell types. By combining GWAS, eQTL mapping, and functional annotation, we elucidated immune mechanisms underlying PsA and further clarified its shared and distinct genetic composition relative to PsO. This multi-layered approach lays the groundwork for developing novel targeted therapies for PsA and facilitates early intervention in PsO patients at elevated risk of PsA.

## Results

### Discovery of new PsA susceptibility loci

To identify PsA risk loci in Europeans, we performed a P-value–weighted meta-analysis of five GWAS, totaling 13,512 PsA cases and 715,639 controls across 8,466,859 bi-allelic autosomal variants^28^ (**Supplementary Data 1**). Our current meta-analysis analysis showed minimal inflation (λ_GC_L=L1.046; LDSC interceptL=L0.9743; **Supplementary** Fig. 1a), suggesting good control for population stratification. Overall, we identified 40 genome-wide significant loci (P<5×10^-8^) outside the MHC region, including 21 novel associations, collectively explaining 23.6% of PsA heritability^29^ (**Fig.1; Supplementary Data 2**). In line with previous studies^30–32^, we detected the strongest associations with PsA in the MHC complex at chromosome 6p22.3 with the lead SNP rs6908205 (OR=1.43, P=3×10⁻²LL) located 33.4 kb upstream of the *HLA-B* gene (**Supplementary Data 2**). This variant exhibited strong linkage disequilibrium (LD) with previously reported lead SNPs associated with PsA, including rs13191343 (r²=0.84, OR=1.41, P=3.8×10⁻²LL) and rs12191877 (r²=0.9, OR=1.42, P=1.4×10⁻²LL). Though genome-wide significant association were observed throughout the MHC, conditional analysis indicated presence of additional two independent loci, with lead SNPs located downstream of the *HLA-A* gene (rs1655901, OR=1.15, P=1.6×10^-90^) and an intronic variant (rs4418214, OR=1.47, P=2.3×10^-266^) in the *MICA* gene (**Supplementary Fig. 1b, Supplementary Data 3**). Outside the MHC, conditional analyses suggested the presence of two independent signals at eight loci (**Supplementary** Fig. 1c-j**, Supplementary Data 4**), which included six at established PsA risk loci and two at novel loci: chromosome 2 (5q15:rs26502, OR=0.96, P=2.3×10-11 and rs2549782, OR=1.042, P=3.03×10^-10^) and chromosome 11 (11q24.3:rs11221249, OR=1.05, P=1.39×10^-12^ and rs11221335, OR=1.047, P=4.7×10^-9^) (**Supplementary Fig. 1c-d**). As an interesting example of our analysis, conditioning on the lead SNP rs2188962 at chromosome 5q31.1, we identified two independent signals spanning the *IL13/IL4* and the *SLC22A5* genes (**Supplementary Fig. 1e)**. Notably, the variant near the *IL13/IL4* gene was protective (rs848, OR=0.91, P=7.7×10^-32^), while those spanning the *SLC22A5* gene conferred increased risk (rs2188962, OR=1.07, P=5.1×10^-23^) for PsA. This is of particular interest given that atopic dermatitis patients treated with dupilumab, an *IL-4R*α inhibitor targeting the IL-13/IL-4 pathway, has been linked to new-onset PsO and PsA^33^; thereby highlighting the potential of GWAS in understanding paradoxical effects of immunomodulatory treatments in genetically predisposed individuals.

**Figure 1.**
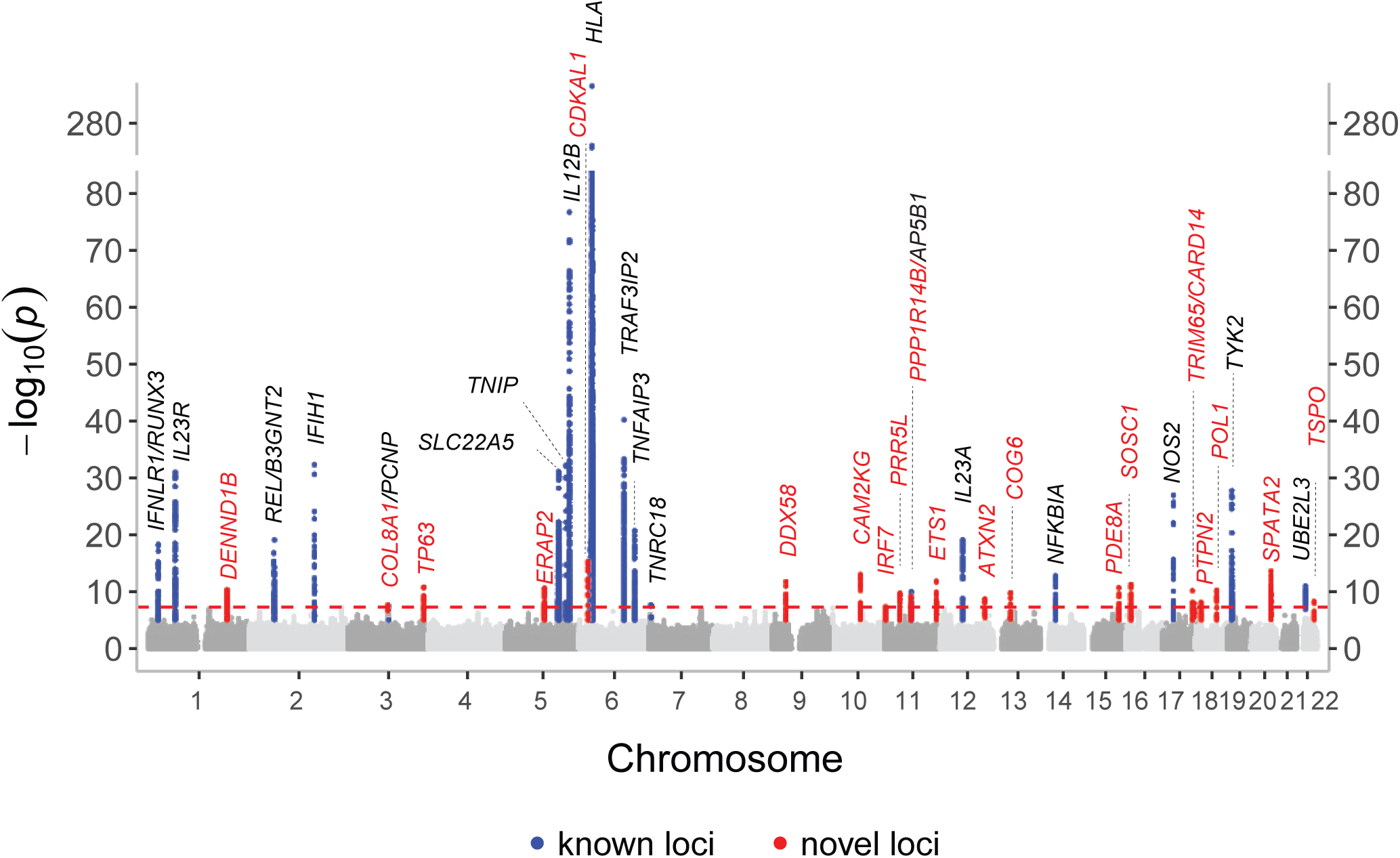
Manhattan plot of genome-wide associations with PsA susceptibility. Manhattan plot showing −log₁₀(P-values) from the meta-analysis of genome-wide associations with PsA susceptibility across individuals of European ancestry. The x-axis represents chromosomal position (hg19), and the y-axis shows statistical significance of association. Red points indicate newly identified loci reaching genome-wide significance (P<5×10⁻L); blue points indicate previously reported loci. The solid red line marks the genome-wide significance threshold. A y-axis break is included between 80 and 275 (dotted line) to improve visualization. Chromosomes 1–22 are alternately shaded for clarity. Gene labels correspond to top-prioritized genes at each locus based on fine-mapping analyses described in the manuscript.

### Refining PsA causal variants using Bayesian fine-mapping approaches

Building on the significant genetic associations in our study, we refined these loci to pinpoint putative causal variants for PsA. To this end, we assessed the causal potential of variants across all non-MHC loci using Bayesian fine-mapping approaches. Within 95% credible sets and a posterior inclusion probability (PP) threshold of >0.95, we identified 623 non-MHC putative causal variants (CVs) using three fine-mapping methods including FINEMAP^34^ (282 CVs), SuSiE^35^ (154 CVs), and SBayesRC^36^ (24 CVs) (**Fig. 2a, Supplementary Data 5**). Incorporating PolyFun priors enhanced precision of CVs prioritization for FINEMAP (236 CVs) and SuSiE (144 CVs). Pathway enrichment analysis^37^ of genes associated with CVs revealed significant enrichment in immune-related and viral response pathways, including measles (FDR=0.00016), JAK-STAT signaling (FDR=0.002), Th1/Th2 cell differentiation (FDR=0.002), RIG-I-like receptor signaling (FDR=0.005), NF-κB signaling (FDR=0.02), and NOD-like receptor signaling (FDR=0.027) (**Supplementary Data 6)**. Consistent with previous findings^38,39^, majority of causal variants were located in intronic (52.1%, nL=L325) or intergenic regions (20.5%, nL=L128), while among the protein-altering variants, 20 missense mutations were distributed across 11 non-MHC PsA loci, including seven within newly identified loci^40^ (**Fig. 2b**). These included deleterious missense variants (CADD score ≥15) in genes known PsA associated loci such as *TRAF3IP2* (rs33980500), *IL23R* (rs11209026), *TYK2* (rs34536443), *SLC35D1* (rs10157422), and *AP5B1* (rs12146493), as well as newly identified loci, including *CARD14* (rs11652075), *DDX58* (rs172217280), and *DENND1B* (rs10494755) (**Supplementary Data 5, Fig. 2c**). While several of these missense variants in *IL23R*, *TRAF3IP2*, *DDX58*, *CARD14*, and *TYK2* were previously identified in PsO GWAS^41^, three other missense variants in *SLC35D1*, *DENND1B*, and *AP5B1* were specific to PsA. One such candidate was the *DENND1B* gene, which encodes DENN domain-containing 1B, a protein involved in regulating T cell receptor (TCR) signaling^42^. In the PsA-weighted locus, we identified one missense variant and four intronic variants within *DENND1B* (**Fig. 2d**). Although a previous GWAS suggested *DENND1B* as a susceptibility locus for psoriasis^43^, limited sample size hindered fine-mapping and causal inference for this gene. Leveraging the increased sample size, our meta-analysis achieved higher resolution and identified causal variants in the *DENND1B* gene associated with PsA.

**Figure 2.**
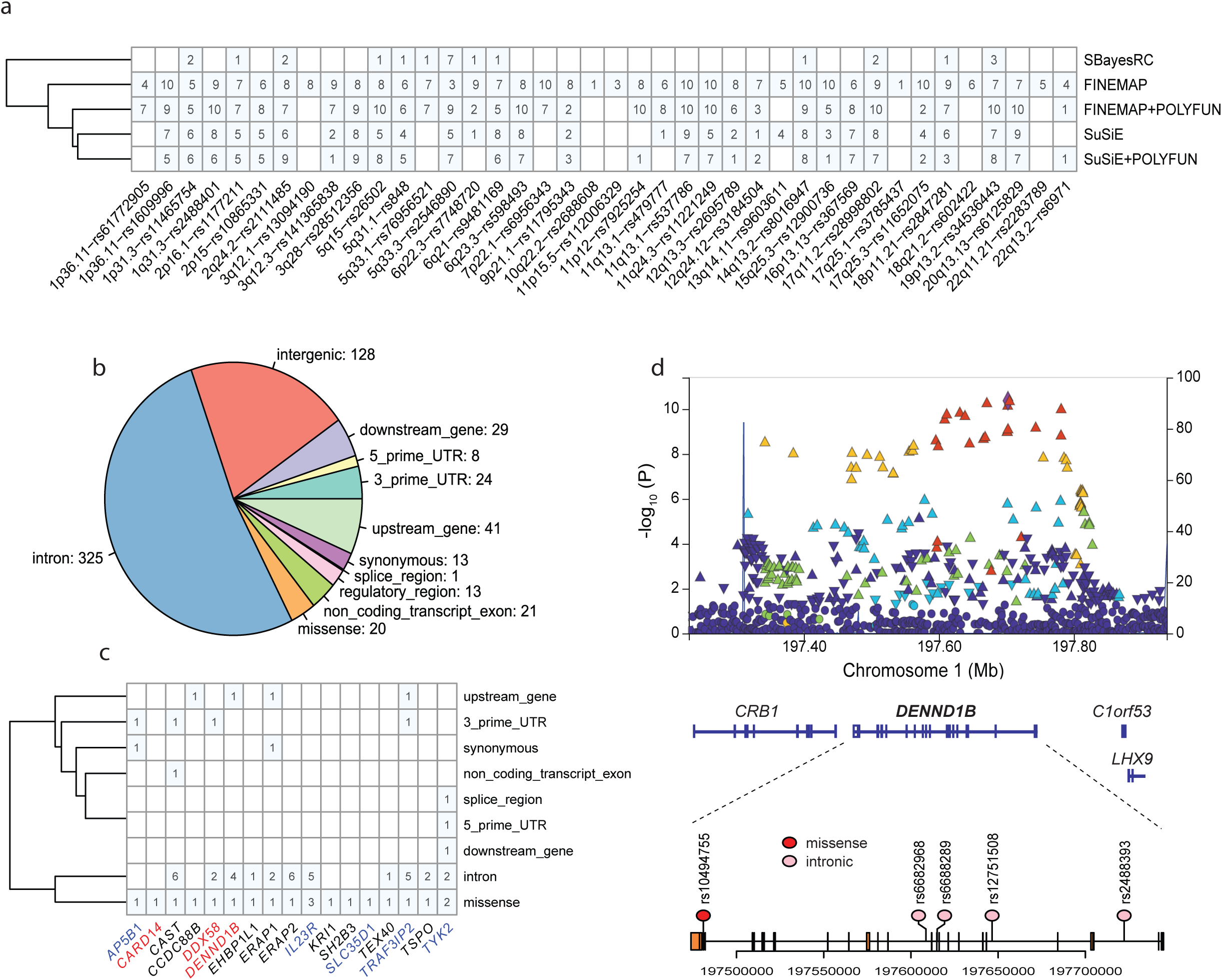
Bayesian fine-mapping of causal variants in PsA. This figure summarizes fine-mapping results across PsA-associated loci. **a** presents a heatmap with fine-mapping tools on the x-axis and cytobands containing lead SNPs on the y-axis. Cells are shaded where multiple causal variants (PP > 0.95) are identified, with counts indicating the number predicted by each tool at each locus. **b** shows a pie chart of the functional consequences of causal variants across all non-MHC PsA loci. **c** displays a heatmap of genes carrying protein-altering missense variants (x-axis) and their predicted consequences (y-axis, based on VEP). Genes with deleterious missense variants (CADD>15) are highlighted in blue if previously known and red if newly identified. **d** features a LocusZoom plot of the *DENND1B* locus, with the upper panel showing –log₁₀(P) values by chromosomal position (Mb) and LD (R²) indicated by color. The lower panel depicts a lollipop plot of DENND1B, with exons as rectangles, red circles for deleterious missense variants, and pink circles for intronic variants.

### Tissues eQTL based prioritization of the candidate genes in PsA

Consistent with prior studies, most PsA-associated variants are non-deleterious^38^, implicating regulatory mechanisms. To quantify the contribution of non-coding variation, we correlated PsA genotypes with gene expression across GTEx tissues^44,45^ and observed enrichment in EBV-transformed lymphocytes, spleen, small intestine (terminal ileum), whole blood, esophagus, and skin, indicating widespread regulatory effects^46^ (**Supplementary Data 7**). Cis-regulated gene expression accounted for 27% of PsA heritability, with tissue-specific contributions from skin (15.7%), whole blood (11.5%), EBV-transformed lymphocytes (11.3%), spleen (8.4%), small intestine (6.8%), and esophagus (5.6%)^47^ (**Supplementary Data 7**).

To pinpoint genes mediating risk, we performed a transcriptome-wide association study (TWAS^48^) using GTEx cis-eQTLs and prioritized SNP–gene pairs with genome-wide significance (P<2×10⁻L, See Methods) and colocalization (COLOC^49^, PP4>0.5). We identified 61 associations across 17 non-MHC PsA loci (**Supplementary Data 8**). We prioritized single genes including *DENND1B*, *ERAP1*, *DDX58*, *RMI2*, *SPATA2, MCAT and TSPO* in novel PsA loci, as well as multiple genes in two novel loci (*11p15.5* and *11q13.1*). Within known loci, we prioritized genes previously associated with PsA, such as *IFNLR1*^50^, *IL23A*^51^, *TYK2*^52^ and *UBE2L3*^53^.

TWAS results can be confounded by LD, where non-causal genes may appear associated due to LD with true causal genes^54^. To address this, we used FOCUS, which accounts for LD structure to prioritize likely causal genes. At PP>0.95 and TWAS significance of P<1.7×10⁻L (See Methods), we identified 41 genes likely causal for PsA-associated risk loci (**Supplementary Data 9**). These included TWAS genes such as *DENND1B*, *ERAP1*, *DDX58*, *RMI2*, *SPATA2, IFNLR1, TYK2 and UBE2L3.* The novel locus at 11p15.5 was further refined to two candidate genes, *PHRF1* and *CDHR5*, while the locus at 11q13.1 was narrowed down to a single gene, *PPP1R14B*. In addition, FOCUS independently prioritized *ERAP2*^55^, *PDE8A*, *SPIRE1*, and *MBD2*^56^ within novel PsA loci, as well as genes previously associated with PsA, including *IL12RB2*^57^ and *B3GNT2*^58^ in known PsA loci.

Next to infer directionality between PsA-associated variants, gene expression, and disease risk, we applied summary-data-based Mendelian randomization (SMR)^59,60^. Integrating PsA GWAS with cis-eQTLs from GTEx v8 (spleen, small intestine, whole blood, esophagus, skin), eQTLGen, and plasma pQTL and mQTL datasets, SMR prioritized 20 genes (GTEx), 29 (eQTLGen), 7 (pQTL), and 42 (mQTL), supporting potential causal regulation (**Supplementary Data 10**). Notably, 27 overlapped with TWAS/FOCUS findings (**Supplementary** Fig. 2). In addition, SMR uniquely implicated *PRR5L* (eQTLGen) at a novel locus (11p12) (**Fig. 3c**), as well as *IL23R* (eQTLGen and mQTL), *IL12B* (pQTL), *REL* (mQTLs) and *NFKBIA* (mQTLs) at known loci, supporting their potential causal role in PsA.

**Figure 3.**
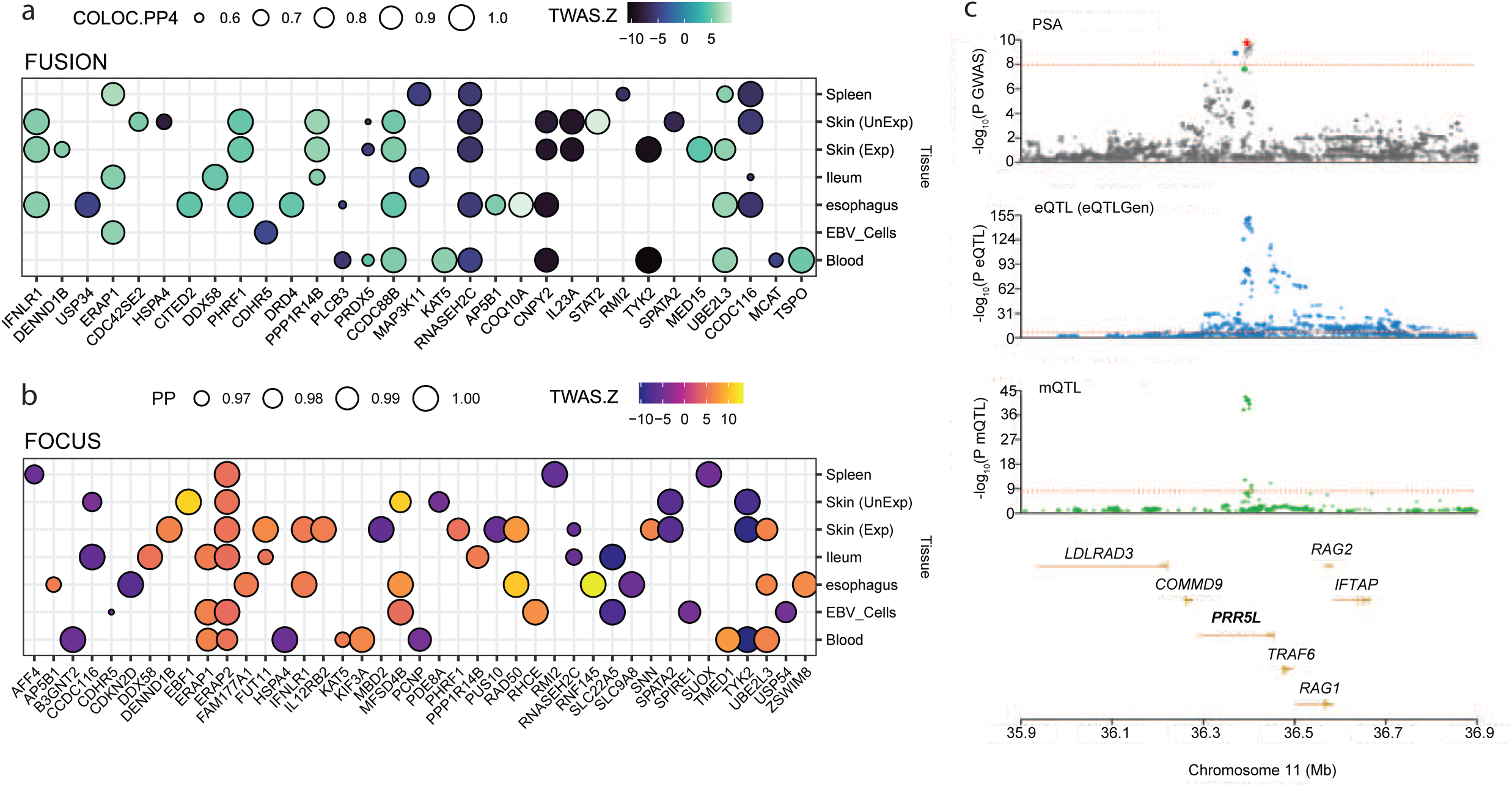
Tissue-based eQTL prioritization of genes in PsA loci. **a** shows a dot plot from the FUSION analysis, with tissues on the y-axis and genes prioritized by FUSION on the x-axis. Dot size represents colocalization probability (PP4), and color indicates the TWAS Z-score. **b** presents a dot plot from the FOCUS analysis, with tissues on the y-axis and genes prioritized by FOCUS on the x-axis. Dot size reflects posterior inclusion probability (PIP), and color represents the TWAS Z-score. **c** shows SMR results for the *PRR5L* gene. The top panel is a LocusZoom plot of the PsA GWAS locus, with lead SNPs for PsA (red), mQTL (green), and eQTL from the eQTLGen cohort (blue). The second panel shows *PRR5L* eQTL results from eQTLGen, and the third shows *PRR5L* mQTL results from the McRae cohort. All three panels display –log₁₀(P) on the y-axis and chromosomal position on the x-axis. The bottom panel shows gene annotations within the locus.

Integrating GWAS data with tissue-specific gene expression profiles, demonstrates the importance of regulatory variation (non-coding variants) in mediating a substantial proportion of PsA risk. By making use of complementary methods, such as TWAS, colocalization, and SMR, we prioritized a set of high-confidence effector genes across both novel and established loci associated with PsA.

### Improved eQTL mapping using pseudobulk analysis and meta-analysis of blood single-cell RNA-Seq data

While tissue-level eQTL mapping has identified numerous candidate genes, interpretation is limited by bulk tissue heterogeneity, which can dilute cell-type-specific effects^61,62^. Cell-type-resolved eQTL analyses improve sensitivity by capturing regulatory variation within relevant cellular contexts^63,64^, but single-cell RNA-seq remains sparse, limiting power^65,66^. To overcome these challenges, we leveraged the OneK1K datasetL and employed a pseudobulk strategy using pre-annotated CELLxGENE cell types to enhance signal-to-noise^67,68^. Re-imputed genotypes enabled single-cell eQTL mapping, identifying 6,989 significant eGenes (±1 Mb of from transcription start sites, P<1×10⁻L, FDR<5%).

Among major immune cell types, CD4⁺ T cells showed the highest number of eGenes (n=4,016), followed by CD8⁺ T cells (n=2,206), NK cells (n=2,141), B cells (n=1,519), monocytes (n=1,062), dendritic cells (n=312), and platelets (n=18) (**Supplementary Data 11**). At finer resolution, enrichment of cis-eQTL signals in specific immune cell subtypes, including CD4⁺ TCM (n=2,840), CD4⁺ naïve cells (n=2,475), CD8⁺ TEM (n=1,706), B naïve cells (n=889), B memory cells (n=602), CD14⁺ monocytes (n=757), and CD16⁺ monocytes (n=623), reflecting the functional heterogeneity underlying in immune cells (**Supplementary** Fig. 3**, Supplementary Data 11**). To further boost the discovery power, we performed meta-analysis (inverse-variance weighted) of OneK1K cohort with the 1M-scBloodNL single-cell cohort^69^. This joint analysis led to an increased number of cis-eQTL, identifying 4,120 eGenes for CD4⁺ T cells, 2,294 for CD8⁺ T cells, 2,205 for NK cells, 1,558 for B cells, 402 for dendritic cells (DCs), and 70 for platelets at the same significance threshold (P<1×10⁻L), illustrating the benefits of cohort-level integration for single-cell eQTL mapping (**Supplementary Data 12, Fig. 4a**).

**Figure 4.**
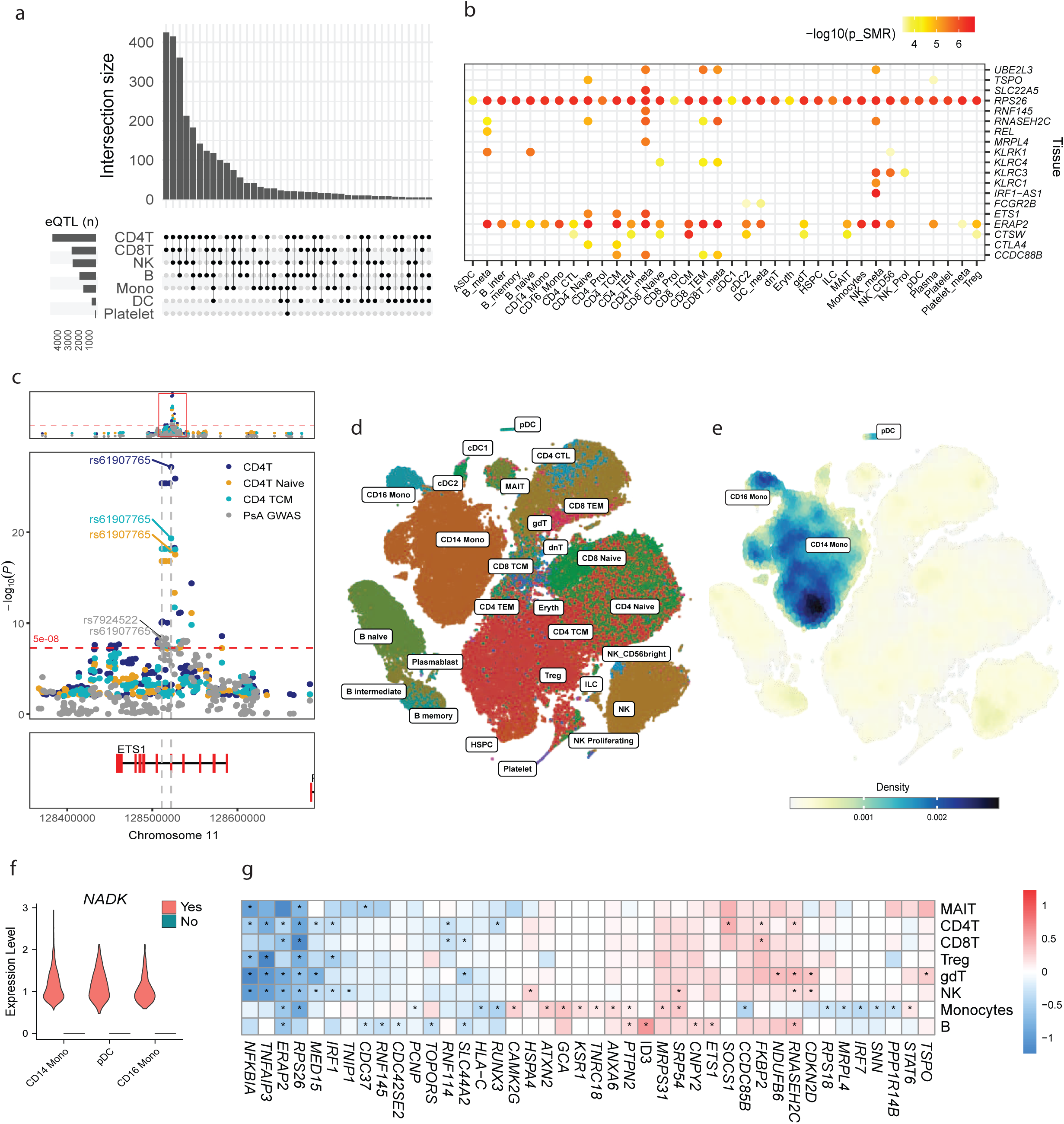
Single-cell eQTL integration and PsA single-cell transcriptomic analysis. **a** UpSet plot showing overlap of significant eGenes (P<1×10⁻L) across major immune cell types in the meta-analyzed OneK1K and 1M-scBloodNL single-cell eQTL datasets. Horizontal bars indicate the number of significant eGenes per cell type; vertical bars show the number of eGenes shared among cell types, as indicated by the filled matrix below. **b** Dot plot of SMR analysis results using single-cell eQTL data, with cell types on the x-axis and SMR-prioritized genes on the y-axis. Dot color reflects −log₁₀(P) values. **c** LocusZoom plot of the ETS1 locus. The top panel shows PsA GWAS −log₁₀(P) values (grey), and the lower panels show eQTL signals in CD4⁺ T cells (blue), naïve CD4⁺ T cells (gold), and central memory CD4⁺ T cells (turquoise). Gene models are displayed with exons in red. **d** t-SNE plot visualizing immune cell clustering in the PsA single-cell dataset. **e** Nebulosa density plot showing enrichment of PsA GWAS-associated cells within the single-cell data. **f** Violin plot comparing NADK expression between GWAS-enriched cells (red) and non-enriched cells (green) CD14+/CD16+ monocytes and pDCs. **g** Heatmap showing log₂ fold change in gene expression between PsA and healthy controls across major immune cell types. Genes are displayed on the x-axis, cell types on the y-axis. Color denotes fold change (blue to red); asterisks indicate differential expression with FDRL<L1%.

Altogether, we have curated a comprehensive resource for cell type–resolved eQTL mapping, which we further harnessed in downstream analyses to fine-map PsA-associated loci at single-cell resolution.

### Cell-type-specific SMR prioritization identifies candidate genes and effector cells in PsA

Next, we applied SMR to prioritize genes using cell type–specific eQTLs and PsA GWAS summary statistics. After adjusting P values for multiple testing within each cell type, we identified several significant associations, including *REL* in B cells (*P*=1.6×10⁻L), *IRF-AS1* in NK cells (*P*=1.7×10⁻L), *SLC22A5* (*P*=5.5×10⁻L) and *RNF145* (*P*=2.4×10⁻L) in CD4⁺ T cells, and *CCDC88B* in both CD4⁺ TCM (*P*=7.1×10⁻L) and CD8⁺ TEM (*P*=4.9×10⁻L) cells. *UBE2L3* was prioritized across CD4⁺ T (*P*=3.1×10⁻L), CD8⁺ TEM (*P*=2.8×10⁻L), and NK (*P*=4.1×10⁻L) cells, while *TSPO* showed associations in CD4⁺ naïve (*P*=1.1×10⁻L) and plasma cells (*P*=3.1×10⁻L) (**Fig. 4b**, **Supplementary Data 13**). Additional genes, including *RNASEH2C*, *CTSW*, *ERAP2*, and *RPS26*, were also prioritized across multiple immune cell populations.

In addition, we identified *ETS1*-prioritized in CD4⁺ naïve (*P*=3.9×10⁻L) and CD4⁺ TCM (*P*=3.1×10⁻L) cells as a novel candidate gene at a newly discovered PsA locus (**Fig. 4c**).

Notably, *ETS1* was not highlighted by tissue based prioritization, underscoring the added value of cellular-resolution eQTL mapping for uncovering disease-relevant candidates. Moreover, our cell type–specific SMR analysis revealed three additional suggestive PsA loci with strong cell-type specificity: 1q23.3, where *FCGR2B* was prioritized in cDC2 cells; 2q33.2, where *CTLA4* expression was detected in CD4⁺ naïve and CD4⁺ TCM cells; and 12p13.2, which showed *KLRK1* expression in B cells, *KLRC4* in CD8⁺ T cells, and *KLRC3/KLRC1* in NK cells (**Fig. 4b, Supplementary Data 13**). Interestingly, Abatacept, a CTLA-4–Ig human fusion protein has already demonstrated clinical efficacy in the treatment of PsA^70^.

Beyond blood-derived immune cells, we also examined cell type–specific cis-eQTLs in keratinocytes, chondrocytes, and osteoclasts; cell types previously implicated in PsA pathogenesis^71–73^ (**Supplementary Data 14**). Although limited by smaller sample sizes, these bulk RNA-seq datasets provided informative, cell-specific insights into regulatory variation. In keratinocytes, SMR identified four significant genes: *ERAP2*, *SLC22A4*, *SLC22A5*, and *SNX32*. These genes were also prioritized in chondrocytes, alongside previously implicated candidates such as *CTSW*, *RPS26*, *NFKBIA*, *RMI2*, and *TSPO*. Notably, chondrocyte cis-eQTLs uniquely prioritized *TRIM65* (*P*=9.1×10⁻L) and *POLI* (*P*=4.2×10⁻L) at two newly identified PsA loci-17q25.1 and 18q21.2, respectively. In osteoclasts, *ERAP2* emerged as a candidate gene.

Collectively, these results advocate for the application of single-cell-resolved QTL-based fine-mapping to improve causal gene inference and suggest that PsA susceptibility is shaped by cell-type-specific contributions from both innate and adaptive immune mechanisms. Furthermore, by capturing both immune and tissue-resident cell types, our approach prioritized candidate genes and their associated effector cells that were not captured by tissue level analysis.

### Polygenic enrichment and differential gene expression reveal disease-specific regulatory signals in PsA immune cells

Previous studies have shown that some eQTLs are stimulation-responsive and highly cell type– specific^74,75^, indicating that analyses limited to single-cell transcriptomic data from homeostatic conditions may overlook critical context-dependent regulatory effects. To address this, we analyzed publicly available single-cell RNA-seq data from peripheral blood of PsA patients, integrated with CITE-seq profiling for precise cell type annotation^76^. Using polygenic regression^77^, we assessed enrichment of immune cell populations associated with non-MHC PsA loci, identifying significant enrichment in innate immune cells including plasmacytoid dendritic cells (pDCs; *P* = 0.0001), CD14⁺ (*P* = 0.0007), and CD16⁺ (*P* = 0.0009) monocytes (**Supplementary Data 15**). Beyond population-level enrichment, polygenic regression revealed enrichment of PsA GWAS signals at the single-cell level: 49.5% of pDCs, 42.7% of CD14⁺ monocytes, and 41.6% of CD16⁺ monocytes showed significant enrichment for genes linked to PsA-associated variants (**Fig. 4d–e**). We classified cells within each population as GWAS-positive (*Padj* < 0.05) or GWAS-negative (*Padj* > 0.05) and performed differential expression analysis between these subgroups (**Supplementary Data 16**). Notably, *NADK* was consistently upregulated in GWAS-positive pDCs and monocytes (**Fig. 4f**), with near absence in GWAS-negative cells, suggesting a potential novel role for *NADK* in PsA pathogenesis and warranting further investigation.

To further investigate disease-specific transcriptional changes, we compared gene expression profiles between PsA patients (n=25) and healthy controls (n=18) across major immune cell populations. At FDR < 0.1, monocytes showed the highest number of differentially expressed genes (n=855; 16.3%) in PsA (**Supplementary Data 17**). Among genes mapped to causal variants (with PP > 0.95 or previously prioritized), *ERAP2*, *NFKBIA*, *TNFAIP3*, *RUNX3*, and *RNF145* that were significantly downregulated across multiple immune subsets in PsA (**Fig. 4g**). *PCNP* and *IRF7* were specifically downregulated only in monocytes. Upregulated genes included *RNASEH2C* (T, B, and NK cells) and *ETS1* (B cells). We also found several genes significantly upregulated in PsA patient blood-*TNRC18* (7p22.1), *ATXN2* (12q24.1), *MRPS31* (13q14.11), and *KSR1* (17q11.2) that have not previously been fine-mapped through colocalization or SMR analyses, with notable expression in monocytes (**Fig. 4g**).

In summary, using disease-specific cell populations, we were able to further fine map causal cell subsets driving the development of PsA and infer putative candidate genes.

### Shared and disease-specific genetic loci between PsO and PsA

PsO precedes the onset of PsA in approximately 15–30% of cases^78^, and more than 90% of individuals with PsA exhibit comorbid PsO^79^, supporting the existence of a shared, at least partially overlapping, genetic susceptibility. Based on this notion, it is reasonable to hypothesize that GWAS in large PsO cohorts may capture signals relevant to PsA, and vice versa.

Using the largest PsO GWAS meta-analysis published to date^41^, we investigated shared and distinct genetic architectures between PsO and PsA. As expected, LDSC analysis revealed a strong positive genetic correlation (rg=1.02, SE=0.032, P=3.22×10⁻²³L) between PsO and PsA. Local genetic correlation analysis revealed significant positive correlations between PsO and PsA at 58.5% (24 out of 41) of PsA-associated loci (See Methods), indicating substantial genetic overlap between the two diseases while also suggesting the presence of disease-specific genetic determinants (**Supplementary Data 18**), as suggested above.

To investigate potential PsA-specific loci, we applied a knockoff-based genome-wide causal inference framework across both PsA and PsO GWAS, leveraging LD from the UK Biobank data (650,000 SNPs)^80^. After harmonizing alleles across summary statistics and reference genotypes, the analysis encompassed 636 LD blocks and 537,193 SNPs. At FDR<0.05, we identified 78 and 45 LD regions harboring putative causal variants for PsO and PsA, respectively (**Supplementary Data 19**). Of the 91 unique LD regions identified, 50% (n=46) were specific to PsO, 14% (n=13) were specific to PsA, and 35% (n = 32) were shared (**Fig. 5a**). Notably, 61% (8/13) of PsA-specific regions were located within 1 Mb of genome-wide significant PsA loci, while the remaining regions showed suggestive associations (P<1×10^-5^). Six of eight putative causal variants were located within known PsA loci including intronic variants in the genes *Lnc-COL8A1* (rs62283827, P=3×10^-6^)*, COG6* (rs7993214, P=1.27×10^-9^)*, PDE8A* (rs12900736, P=1.95×10^-11^)*, PTPN2* (rs2847281, P=6.87×10^-9^), and a missense variant in *SPATA2* (rs495337, P=2.3×10^-13^). Two other causal variants were located on novel PsA risk loci 7p22.1 (rs11770660, P=3.37×10^-8^) and 22q13.2 (rs138922, P=6.29×10^-7^), which fine-mapped to the genes *TNRC18* and *TSPO*.

**Figure 5.**
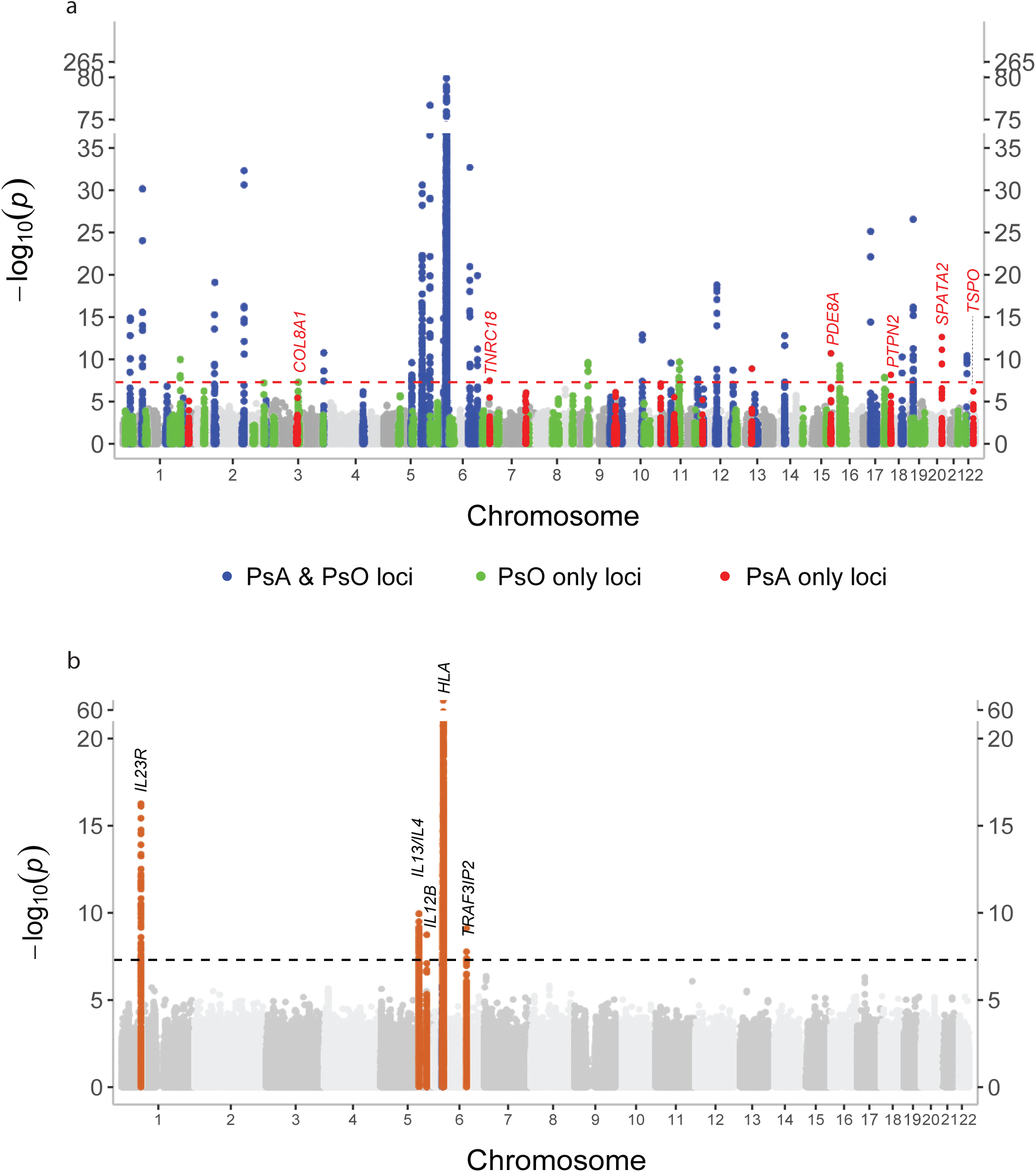
Genome-wide association of PsA-specific signals using causal inference and conditional analysis. **a** Manhattan plot of −log₁₀(P) values from the GhostKnockoffGWAS model in PsA. The x-axis shows chromosomal position (hg19), and the y-axis reflects statistical significance. Red points mark PsA-specific causal variants; green points indicate PsO-specific variants. The horizontal red line marks genome-wide significance (PL<L5L×L10⁻L). A y-axis break is shown between 80 and 265 to improve visualization. Chromosomes 1–22 are alternately shaded, and gene labels are shown for genome-wide significant PsA loci. **b** Manhattan plot of −log₁₀(P) values from multi-trait conditional and joint analysis comparing PsA and PsO. Red points indicate PsA-specific loci reaching genome-wide significance. A y-axis break is shown between 20 and 60. Cytoband labels mark significant PsA-specific loci.

To assess whether PsA-associated loci exert effects independent of PsO, we applied multi-trait conditional and joint analysis (mtCOJO^81^).. This analysis revealed PsA-specific signal at the MHC locus (rs34902309, OR = 1.29, P = 2.8×10⁻L¹), located near the *HLA-B* gene. Outside the MHC region, four additional loci demonstrated PsA-specific associations: 1p31.3 (rs6688383, OR=1.06, P=5.5×10⁻¹L), 5q31.1 (rs12521097, OR=1.05, P=1.1×10⁻¹L), 5q33.3 (rs2546890, OR=1.04, P=1.8×10⁻L), and 6q21 (rs13199291, OR=1.09, P=7.8×10⁻¹L) (**Supplementary Data 20, Fig. 5b**). Notably, both the PsA specific MHC region and the IL23R-containing 1p31.3 loci have been previously implicated in genetic differentiation between PsO and PsA^18^. In addition, our conditional analysis, suggested PsA-specific genetic signature at several known loci harboring immune-related genes, including *P4HA2*, *IL12B*, and *TRAF3IP2*, further reinforcing their PsO-independent relevance to PsA pathogenesis.

Taken together, these results demonstrate that while increasing GWAS sample size improves power to detect causal variants, fine phenotypic resolution is critical for uncovering disease-specific associations. Thus, broadly defined case groups such as combined PsO/PsA cohorts may obscure signals specific to PsA. Our findings highlight the value of refined PsA-focused analyses to disentangle shared from distinct genetic contributions.

## Discussion

Previous GWAS studies have identified numerous loci associated with PsA, yet pinpointing causal genes and variants remained difficult often hindered by limited cohort sizes. Accordingly, larger GWAS in PsO have demonstrated that increased sample size enhances both locus discovery and fine-mapping resolution^41^. In contrast, our findings underscore that in addition to increased statistical power, clinical phenotyping is essential for resolving causal signals. The current study identified 40 loci outside the MHC region, of which 21 were new and harbored putative genes encoding for potential therapeutic targets (**Supplementary Data 21**). These included 3 deleterious missense variants in *SLC35D1*, *AP5B1, and DENND1B* genes, which were specific to PsA. Interestingly, although the *DENND1B*-harboring locus was previously reported in a larger PsO GWAS (with three times the sample size), such precise variant-level resolution was not attained.

Using tissue-informed TWAS, we prioritized genes encoding therapeutic targets that are already modulated by approved PsA treatments. These include *IL23A*, *IL23R*, *IL12B*, and *TYK2*, which are targeted by guselkumab, risankizumab, and ustekinumab-all of which are approved for PsA^82–84^, as well as deucravacitinib, which targets TYK2 and is approved for PsO but currently under investigation for PsA^85^. In addition, we report 22q13.2 as a novel genome-wide significant PsA locus, not identified for PsO. At this locus, TWAS highlighted the *TSPO* gene that encodes for the mitochondrial translocator protein (rs80411, PL=L3.2L×L10⁻L, PPL=L0.997), which has been previously associated with monocyte, platelet, and eosinophil counts^86^. *TSPO* is highly expressed in activated macrophages and fibroblast-like synoviocytes in arthritic joints^87^ and has been used as an imaging biomarker of synovial inflammation in rheumatoid arthritis^88^. Its prioritized role in PsA suggests a broader involvement in joint-infiltrating immune and stromal cell activation, pointing to a previously unrecognized link to PsO. Transcriptome-wide SMR analysis further prioritized *PRR5L*, a key component of the TORC2 complex that modulates mTOR signaling^89^ (P=1.29×10⁻L). Notably, aberrant mTOR pathway activation has been implicated in driving synovial fibroblast proliferation and chronic inflammation in PsA^90^, highlighting *PRR5L* as a promising candidate for further investigation in psoriatic disease^91^.

While bulk eQTLs offer broad regulatory insights, cell-type–specific eQTLs are essential for dissecting disease-relevant mechanisms^65^. Existing single-cell datasets (e.g., OneK1K, 1M-scBloodNL) lack effect size estimates required for integrative analyses like SMR. To overcome this, we reprocessed OneK1K and meta-analyzed it with 1M-scBloodNL, generating high-resolution eQTL maps across immune cell types. These enhanced datasets enabled cell-specific gene prioritization for PsA and represent a valuable resource for future fine-mapping efforts.

Taking advantage of these newly curated datasets, we prioritized *ETS1* (E26 transformation-specific sequence 1) at 11q24.3, a transcription factor previously implicated in PsO^92^ but newly associated with PsA in our study. We identified the expression of *ETS1* in CD4⁺ naïve and CD4⁺ TCM cells, which is consistent with its role in mediating T cell differentiation^93^. Accordingly, *ETS1* hypomorphic mice show impaired Th1 differentiation and enhanced Th17 responses, suggesting a plausible role for ETS1 in PsA pathogenesis^94^. Among the PsA-associated stromal cell types, such as keratinocytes, chondrocytes, and osteoclasts, we prioritized the gene *ERAP2*, which has been previously linked to PsA risk, particularly among HLA-B27-negative individuals in a Romanian cohort^95^.

Utilizing PsA-specific single-cell RNA-seq data from blood, we observed significant non-MHC GWAS signal enrichment in pDCs and CD14⁺/CD16⁺ monocytes. Notably, all three cell types exhibited elevated expression of *NADK*. Nicotinamide adenine dinucleotide kinase (NADK) is a key enzyme that phosphorylates NAD⁺ to produce NADP(H), molecules critical for antioxidant defense and biosynthetic metabolism^96^. Importantly, murine studies have shown that limiting NAD⁺ availability impairs Th17 cell differentiation and function, suggesting that NADK activity may support immune modulating role in PsA^97^. These findings call for further investigation into the role of NADK in PsA.

Lastly, we examined the shared and distinct genetic architectures of PsA and PsO. Despite the strong genetic correlation between these diseases, we initially expected minimal differentiation. However, our analyses uncovered notable differences in the profiles of putative causal variants. By leveraging fully harmonized GWAS summary statistics, we identified PsA-specific signals at loci including *COL18A1*, *TNRC18*, *PDE8A*, *PTPN2*, *SPATA2*, and *TSPO*. Although these loci have been previously implicated in PsO, the causal variants we pinpointed appear specific to PsA, suggesting that these associations may be driven predominantly by the PsA subset within PsO cohorts. In support of this notion, an independent Italian study reported a stronger effect size for *COL8A1* in PsA (ORL=L2.24) compared to PsO (ORL=L1.85)^98^. Furthermore, using mtCOJO, we identified additional PsA-specific loci, including the MHC, 1p31.3, 5q31.1, 5q33.3, and 6q21. Notably, four of these regions (MHC, 1p31.3, 5q31.1, and 5q33.3) harbored multiple independent signals based on GCTA-COJO analysis. For example, at 1p31.3, we detected a protective signal near *IL23R* and a pathogenic signal near *C1orf141*. Conditioning on PsO attenuated the IL23R-associated signal, while the *C1orf141* signal remained significant, suggesting PsA-associated specificity. A similar pattern was observed at 5q31.1, where signals mapped to *IL13/IL4* (protective) and *P4HA2* (pathogenic) genes. Only the *P4HA2* association persisted after conditioning, consistent with prior PsA reports^99^. At 5q33.3, we observed protective and pathogenic signals near the *IL12B* and the *UBLCP1* genes, respectively, with only the *IL12B* association remaining significant post-conditioning, further highlighting PsA-specific genetic architecture.

Collectively, our integrative analyses reveal the unique molecular mechanisms driving PsA, distinguishing it from the overlapping genetic architecture of PsO. By identifying putative PsA-specific causal variants and candidate genes, we lay the groundwork for targeted therapeutic development and provide a comprehensive resource to advance future fine-mapping and functional studies aimed at elucidating disease pathogenesis.

## Methods

### GWAS meta-analysis and conditional analysis

We performed a genome-wide meta-analysis across five cohorts of European ancestry: the Spanish cohort, North American cohort, FinnGen, UK Biobank, and the Million Veteran Program (MVP)^17,18,100,101^. To account for methodological differences in the generation of GWAS summary statistics across cohorts, we employed the p-value weighted meta-analysis scheme as implemented in the METAL^28^ software. This approach was chosen because summary statistics for the Spanish, North American, and UK Biobank cohorts were derived using linear regression models, whereas the FinnGen and MVP cohorts utilized linear mixed models to adjust for population structure and relatedness. All GWAS summary statistics were harmonized to a common set of variants with minor allele frequency (MAF) ≥ 1% in the European population from the 1000 Genomes Project^102^. This harmonization step ensured alignment of effect alleles, removal of strand ambiguous SNPs, and exclusion of variants with mismatched alleles or population-specific inconsistencies. For each dataset, we calculated the effective sample size using the formula: Neff = 4 × Ncases × Ncontrols / (Ncases + Ncontrols), to appropriately weight each study in the meta-analysis. During quality control, we observed moderate genomic inflation (λ > 1.05) in the FinnGen and MVP datasets. To mitigate potential confounding from residual population stratification or cryptic relatedness, we applied genomic control (GC) correction to these cohorts prior to their inclusion in the meta-analysis^103^. Following meta-analysis, we further excluded SNPs exhibiting allele frequency deviations greater than 20% compared to the European reference panel from the 1000 Genomes Project. This step was implemented to reduce artifacts potentially arising from genotyping errors, strand mismatches, or population-specific discrepancies.

Initially, meta-analysis identified 44 independent genomic loci exceeding the genome-wide significance threshold (P<5×10^-8^) including HLA, as determined using the FUMA (Functional Mapping and Annotation) pipeline^46^. To evaluate heterogeneity in effect sizes across cohorts and reduce the likelihood of false positives, we applied a Bonferroni correction based on the number of significant loci^41^. Specifically, we adopted a heterogeneity p-value threshold of P<1.14×10^-3^ (0.05/44), as previously described, to identify and exclude associations with substantial between-cohort variability that may not represent true underlying genetic signals. This resulted in 41 genome-wide significant PsA loci.

To identify conditionally independent association signals within each genome-wide significant locus, we conducted a stepwise conditional analysis using GCTA-COJO (v1.93.2beta)^104^. We first defined genomic regions by extending ±1.5 Mb around lead SNPs identified by FUMA and merged overlapping intervals using BEDTools to delineate 37 non-overlapping loci^105^. For each region, we extracted the corresponding summary statistics and generated locus-specific genotype reference panels using the European subset of the 1000 Genomes Project (n = 503) and OneK1K^106^ datasets (n = 917), processed via PLINK2 (v2.0). Summary statistics were formatted according to COJO requirements, including SNP ID, effect and non-effect alleles, allele frequency, effect size, standard error, and p-value^107^.

The analysis began by identifying the most significant SNP within each region and conditioning on it to detect additional independent signals. This process was iteratively repeated, adding newly identified SNPs to the conditioning set and re-estimating conditional associations. We retained SNPs that remained genome-wide significant (P<5×10⁻L), and were located at least 500 kb from any previously conditioned SNP. This locus-specific approach to LD modeling enabled precise detection of independent signals. The final set of conditionally independent variants was compiled across all loci.

### Bayesian fine-mapping and functional annotation of PsA loci

To identify candidate causal variants at genome-wide significant loci, we applied multiple complementary Bayesian fine-mapping approaches, including Sum of Single Effects (SuSiE)^108^, FINEMAP^34^, and SBayesRC^36^. SuSiE, implemented in R, uses sparse regression modeling to estimate posterior inclusion probabilities (PIP) and define credible sets of likely causal variants. FINEMAP performs a shotgun stochastic search using summary statistics and linkage disequilibrium (LD) matrices to model multi-SNP causal configurations. SBayesRC, implemented in the GCTB software^109^, applies Bayesian multiple regression to GWAS summary statistics while incorporating functional annotations to improve causal inference. Fine-mapping analyses were performed within ±1.5 Mb windows around each lead SNP using an ancestry-matched LD reference panel. We constructed 95% credible sets for each method, and performed fine mapping using with and without functional priors from PolyFun^110^ where appropriate to improve resolution. To functionally annotate variants within credible sets, we used the Ensembl Variant Effect Predictor^40^ (VEP v109) to assess coding and regulatory consequences, including predicted missense and splice-altering effects. Variant deleteriousness was assessed using Combined Annotation Dependent Depletion (CADD) scores^111^. To explore the biological relevance of fine-mapped loci, we conducted pathway enrichment and protein– protein interaction analyses using the STRING database^37^ (v12.0) and g:Profiler, referencing Gene Ontology (GO)^112^ and KEGG pathway^113^ databases. All enrichment analyses were FDR-adjusted to account for multiple testing.

### Tissue based gene prioritization of PsA loci

To identify tissues in which PsA-associated variants are enriched, we performed tissue-specific gene expression enrichment analysis using FUMA^46^. Genome-wide significant SNPs (P < 5 × 10⁻L) were mapped to candidate genes via FUMA’s SNP2GENE function, which integrates positional mapping, eQTL mapping, and chromatin interaction data. These genes were then tested for enrichment in tissue-specific expression profiles using the GTEx v8 dataset^45^. FUMA evaluates overexpression relative to a background distribution and applies false discovery rate (FDR) correction across all GTEx tissues. To quantify the contribution of cis-regulated gene expression to PsA heritability, we applied mediated expression score regression (MESC) using eQTL data from the GTEx tissues identified as enriched by FUMA^47^.

For gene prioritization, we performed a transcriptome-wide association study (TWAS) using the FUSION pipeline^54^. PsA GWAS summary statistics were harmonized and aligned to the 1000 Genomes Phase 3 European reference panel for LD estimation. Gene expression prediction models trained on GTEx v8 data (including sun-exposed and non–sun-exposed skin, whole blood, EBV-transformed lymphocytes, spleen, terminal ileum, and esophagus) were used. TWAS association testing was conducted using the FUSION.assoc_test.R script, incorporating LD to account for SNP correlations. Associations within non-MHC genome-wide significant loci were retained using the GenomicRanges R package^114^. Across tissues, we identified 2,459 genes–SNP pairs. Bonferroni correction (P<2 × 10⁻L; 0.05/2459) was applied to determine significance. To evaluate whether TWAS signals were driven by shared causal variants, we conducted colocalization analysis using COLOC^49^ within FUSION. For each gene, COLOC estimated the posterior probability that a single causal variant explained both the GWAS and eQTL signals, within ±500 kb of the gene (via FUSION.post_process.R). Genes with posterior probability (PP4) > 0.5 were considered to have evidence of a shared causal signal.

Because TWAS signals can be confounded by LD, we further fine-mapped candidate genes using FOCUS (Fine-mapping of CaUsal Gene Sets)^54^. FOCUS jointly models TWAS statistics, accounting for LD and correlations among expression models, to estimate posterior inclusion probabilities (PIPs) and construct 90% credible gene sets. FOCUS was applied to each non-MHC genome-wide significant locus, across enriched tissues, using the same GWAS data, gene expression weights, and LD reference panel as in FUSION. This analysis yielded 2,921 significant tissue–SNP–gene combinations. A Bonferroni threshold (P<1.7×10⁻L; 0.05/2921) was used to define significance. Only pairs with PIP ≥ 0.95 were retained. All visualizations were generated using the ggplot2 R package^115^.

To assess whether PsA-associated variants influence disease risk via gene expression (i.e., mediation) rather than through pleiotropy, we applied Summary-data-based Mendelian Randomization (SMR) using the SMR portal^60^. SMR integrates GWAS and eQTL summary statistics to test whether genetically regulated gene expression mediates GWAS associations. We uploaded PsA GWAS summary statistics to the portal using default settings. SMR was conducted separately for GTEx tissues enriched by FUMA (sun-exposed and non–sun-exposed skin, whole blood, spleen, terminal ileum, esophagus), and external datasets including eQTLGen (blood), mQTL (blood), and pQTL (INTERVAL^116^, FENLAND^117^, SCALLOP^118^). The UK Biobank reference panel^119^ (provided by default in the portal) was used for LD estimation. Bonferroni-adjusted significance thresholds were applied to each dataset as follows: Whole Blood (P<1×10⁻L), Skin (Sun-Exposed) (P<8.5×10⁻L), Skin (Not Sun-Exposed) (P<9.5×10⁻L), Esophagus (P<9.6×10⁻L), Terminal Ileum (P<3.5×10⁻L), Spleen (P<1.7×10⁻L), eQTLGen (P<4.5 ×10⁻L), FENLAND (P<3.2×10⁻L), INTERVAL (P< 8.1×10⁻L), SCALLOP (P<7×10⁻L), and mQTL (P<3.8×10⁻L). Only SNP–gene pairs where the cis-eQTL association reached genome-wide significance (P<5×10⁻L) were retained to reduce false positives due to weak instruments.

### cis-eQTL mapping in the Onek1k cohort and integration with external datasets

The Onek1k project^106^ provides a rich single-cell eQTL resource based on blood samples from 982 donors, offering valuable insights into gene regulation at the cellular level. Although summary statistics are publicly accessible, they are not directly compatible with many Mendelian randomization (MR) frameworks^47^, which typically require individual-level genotype data or harmonized variant representations. Moreover, discrepancies exist between the cell type annotations in the Onek1k dataset and those provided by large-scale, harmonized single-cell reference atlases such as Human Cell Atlas (HCA)^120^, reflecting expected differences in annotation methods and reference hierarchies. To address these limitations and enhance compatibility, we re-processed the raw genotyping data from the Onek1k cohort.

We obtained Illumina IDAT files from the NCBI GEO repository^121^ and processed them using GenomeStudio 2.0. One sample showing a discrepancy between reported and genotypic sex was excluded. Ancestry was assessed with KING v2^122^ using the 1000 Genomes Project as a reference panel, and one non-European ancestry outlier was removed. Principal component analysis (PCA) was then performed iteratively with PLINK v2, and samples identified as outliers based on the first 10 PCs were excluded after manual inspection, resulting in a final dataset of 917 samples and 508,841 high-quality bi-allelic SNPs (MAF > 1%, call rate > 95%, HWE P < 1×10⁻L). To increase marker density for downstream analyses, genotype imputation was performed using the TOPMed reference panel^123^. Post-imputation quality control in PLINK2 excluded variants with imputation quality R² < 0.8, MAF < 1%, or HWE P < 1×10⁻L. This resulted in a high-quality imputed dataset of 7,651,534 SNPs across 917 individuals, suitable for single-cell eQTL analyses and Mendelian randomization studies.

We retrieved a processed single-cell gene expression dataset in H5AD format from the CELLxGENE portal^68^, which included cell-level annotations based on the predicted. celltype. l2 field. To generate pseudo-bulk gene expression profiles, raw UMI counts were aggregated across individual donors and both major and fine-resolution immune cell populations using the AggregateExpression function from the Seurat R package^124^. Major immune cell categories were defined by collapsing biologically related subtypes as follows: B cells (B intermediate, B memory, B naive, Plasmablast), CD4⁺ T cells (CD4 CTL, CD4 naive, CD4 proliferating, CD4 TCM, CD4 TEM), CD8⁺ T cells (CD8 naive, CD8 proliferating, CD8 TCM, CD8 TEM), monocytes (CD14⁺ and CD16⁺), NK cells (NK, NK proliferating, NK_CD56bright), and dendritic cells (cDC1, cDC2, pDC, ASDC). To reduce noise from low-abundance transcripts, genes with fewer than 5 UMI counts in at least 20 samples were excluded. The aggregated count data were normalized using the trimmed mean of M-values (TMM) method implemented in the edgeR package^125^, followed by log-transformation to obtain log-counts per million (logCPM). To adjust for potential technical and biological confounders such as donor pool and age, we applied linear modeling to regress out these covariates. The resulting residuals were then subjected to gene-wise inverse-rank normalization to ensure comparability across samples and approximate a normal distribution of expression values^126^.

We conducted cell type–specific cis-eQTL analysis by correlating genotype with gene expression using the MatrixEQTL (linear model) R package^127^. The analysis model included sex as a covariate and the top 10 genotype principal components to account for population structure. A genetic relatedness matrix was computed using EMMAX^128^ and supplied to MatrixEQTL as a covariance matrix to correct for distant relatedness among donors. We identified cis-eQTLs within ±1 Mb of a gene’s transcription start, and associations were considered statistically significant at a FDR of <5% and P<1×10^-5^. For cross-study meta-analysis, we retrieved eQTL summary statistics for major immune cell types from the sc-eQTLGen consortium (1M-scBloodNL, untreated)^69^. Genomic coordinates were converted from hg19 to hg38 using the rtracklayer package in R. Meta-analysis between Onek1k and sc-eQTLGen was conducted using the inverse-variance weighted (IVW) method.

Additional cis-eQTL summary statistics for tissue-resident cells-keratinocytes, chondrocytes, and osteoclasts were sourced from publicly available datasets^71,76,129^. These datasets were harmonized using Onek1k and 1000 Genomes references to correct for strand flips, allele mismatches, and genome build differences.

All cis-eQTL summary statistics were converted to the binary effect size summary data (BESD) format for integration into SMR software (v1.3.1), using Onek1k as the reference panel. To maintain consistency with tissue-level eQTL analyses, we excluded associations within the MHC region. Only genome-wide significant eQTLs were considered for gene prioritization in downstream MR analyses. Bonferroni correction was applied separately per cell type. Plotting was performed using ComplexUpset^130^ and ggplot2 R packages.

### GWAS enrichment analysis in PsA single-cell blood transcriptome

We obtained publicly available scRNA-seq data from peripheral blood samples of 25 individuals diagnosed with psoriatic arthritis (PsA) and 18 healthy controls^76^. Quality control was performed using the Seurat package. To retain high-quality cells, we excluded those with fewer than 1,000 or more than 20,000 unique molecular identifiers (UMIs), as low UMI counts may indicate empty droplets or dying cells, while high UMI counts can suggest doublets or multiplets. Cells expressing fewer than 500 or more than 3,000 genes were removed to eliminate low-complexity or overly complex profiles, which can reflect poor-quality cells or technical artifacts. Additionally, cells with >10% mitochondrial gene expression were excluded, as this may indicate cellular stress or apoptosis. After filtering, 71,016 high-quality PsA cells and 71,812 control cells were retained for downstream analysis.

To identify trait-relevant cell types, we used scPagwas^77^, a pathway-level polygenic regression method that integrates GWAS summary statistics with scRNA-seq data. scPagwas maps GWAS variants to genes through curated pathway annotations and models the relationship between genetic risk and single-cell gene expression. In our analysis, we used PsA GWAS summary statistics alongside the filtered PsA single-cell dataset. Variants in high linkage disequilibrium (LD, R² > 0.8) were pruned using PLINK2, and variants in the MHC region (chr6:25–35 Mb) were excluded due to their disproportionate effect on immune-related signals. A polygenic regression model was then used to assign polygenic scores to individual cells, allowing the identification of cell types enriched for trait-associated signals. The analysis was conducted using the scPagwas_main2 function with default parameters. Statistical significance was evaluated using a bootstrapping procedure, and enrichments with P<1.1×10⁻L were considered significant. Cell populations with FDR < 1% were considered trait-relevant. Cells were labeled as GWAS-enriched or non-enriched based on this threshold, and differential expression analysis between these groups was conducted using the Wilcoxon test implemented in Seurat. Visualization was performed using the SCpubr R package^131^.

For differential gene expression analysis between PsA and healthy controls, we aggregated cells into major populations and created pseudobulk profiles for each sample within each cell type. Differential expression was performed separately for each population. Genes with fewer than five counts across samples or expressed in fewer than 10% of cells were excluded. Normalization was performed using the computeSumFactors() function from the scran package^132^ to estimate cell-specific size factors and correct for sequencing depth. Differential expression was then assessed using the DESeq2 package^133^. Gene-wise dispersion was estimated using the glmGamPoi fitting method, and likelihood ratio tests were used to compare the full model to a reduced model (∼1), identifying genes with significant differences between groups. Only genes located within genome-wide significant non-MHC PsA loci were retained. P-values were adjusted using false discovery rate (FDR) correction, and the results were visualized using the pheatmap R package^134^.

### Correlation, causal, and conditional analysis between PsA and PsO

Summary statistics for PsO GWAS were downloaded from the GWAS Catalog. The data were harmonized to the 1000 Genomes Project reference panel (hg19), with quality control steps including strand alignment, allele matching, and restriction to biallelic SNPs. This resulted in a set of 8,328,763 high-quality variants for downstream analysis. Global genetic correlation between PsA and PsO was assessed using the LDSC (LD Score Regression) software, which estimates genome-wide heritability and genetic correlation from GWAS summary statistics while accounting for LD structure^135^. Specifically, munge_sumstats.py was used to format the summary statistics using the w_hm3.snplist, excluding the MHC region. Genetic correlation was then estimated using ldsc.py with default parameters across 1,146,450 common SNPs.

Local genetic correlation was calculated using the LAVA (Local Analysis of [co]Variant Association) R package^136^, which partitions the genome into LD blocks and models local genetic covariance between traits. Predefined European LD blocks (n=2,495) were downloaded from the LAVA GitHub repository. To correct for potential sample overlap between studies, genetic covariance between PsA and PsO was estimated using LDSC. Each LD block was first tested for univariate genetic signal in both PsA and PsO datasets. Blocks showing suggestive association (P<1×10⁻L) in either trait were then tested for bivariate local genetic correlation. Statistical significance was determined using a Bonferroni-corrected threshold of P<2.03×10⁻L (0.05/2,459).

To identify likely causal variants in PsA and PsO GWAS datasets, we employed GhostKnockoffGWAS^80^, a recently developed fine-mapping framework that identifies sets of variants carrying non-redundant information about the phenotype across the genome. This method leverages a knockoff-based inference strategy to perform genome-wide conditional independence testing using only GWAS summary statistics, enabling robust detection of putative causal variants while accounting for linkage disequilibrium and controlling the false discovery rate (FDR).To mitigate potential biases arising from allele or SNP mismatches, we restricted the analysis to SNPs (N=8,280,251) that had identical reference and alternate alleles in both the PsA and PsO GWAS summary statistics. Z-scores for each variant were calculated by dividing the effect size (beta) by its standard error. Causal variants were identified separately for each GWAS using a matched SNP reference panel (UK Biobank; SNPs = 537,193). Variants were deemed causal if they passed an FDR threshold of < 5%. Functional annotation of the identified causal variants was performed using g:Profiler^137^.

To evaluate whether PsA-associated loci exert effects independent of PsO, we employed multi-trait conditional and joint analysis (mtCOJO) within the COJO framework^81^. This approach conditions PsA summary statistics on PsO GWAS to account for shared polygenic background, enabling the identification of trait-specific effects. For the execution of mtCOJO, the PsA and PsO GWAS summary statistics were converted into the COJO format (.ma files). The 1000 Genomes European reference data in PLINK format, along with the European LD reference (eur_w_ld_chr) provided by LDSC software, were used for LD estimation. The analysis included 8,280,239 bi-allelic variants that were common across the PsA, PsO, and 1000 Genomes reference datasets. Manhattan plots for both the causal and conditional analyses were generated using the topr R package.

## Data Availability

The GWAS summary statistics were obtained from the GWAS Catalog, including the Spanish and North American cohort (GCST007043), UK cohort (GCST90243956), and the Million Veteran Program (GCST90476191). Finnish GWAS summary statistics were retrieved from the FinnGen database (https://public-metaresults-fg-ukbb.finngen.fi/pheno/M13_PSORIARTH). Individual-level reference data from the 1000 Genomes Project were accessed through the NYGC 1000 Genomes portal (https://ftp.1000genomes.ebi.ac.uk/vol1/ftp/data_collections/1000G_2504_high_coverage/working/20201028_3202_raw_GT_with_annot/) for conditional and causal variant analyses. FUMA/MAGMA analyses were conducted using the FUMA platform (https://fuma.ctglab.nl/). Summary-level Mendelian randomization was performed using the SMR portal (https://yanglab.westlake.edu.cn/smr-portal/). Single-cell gene expression data for the OneK1K cohort were obtained from the CELLxGENE portal (https://cellxgene.cziscience.com/collections/436154da-bcf1-4130-9c8b-120ff9a888f2), and the corresponding individual-level genotype data were retrieved from NCBI GEO (GSE196830). eQTL summary statistics for immune cell populations from the 1M-scBloodNL single-cell cohort were obtained from the sc-eQTLGen portal (https://eqtlgen.org/sc/datasets/1m-scbloodnl-eqtls.html). Single-cell transcriptomic data for Psoriatic Arthritis patients were accessed from NCBI GEO (GSE194315). Finally, psoriasis GWAS summary statistics were obtained from the GWAS Catalog (GCST90472771).

