## Supplementary Figures for "Genome-wide meta-analysis and integrative fine-mapping identify novel susceptibility loci and effector genes in psoriatic arthritis"

for

*Gupta et al.*

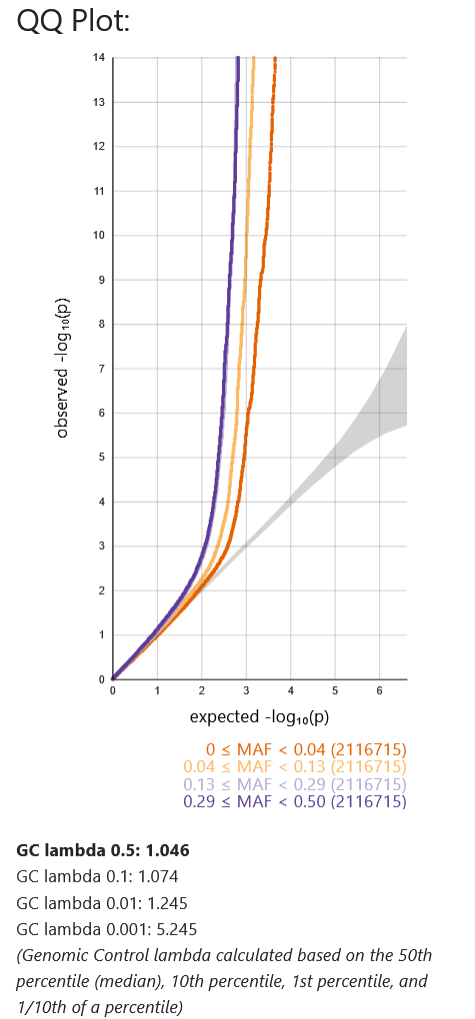

**Supplementary Fig. 1a.** QQ plot Genomic control lambda calculated based on the 50^th^ percentile (median.

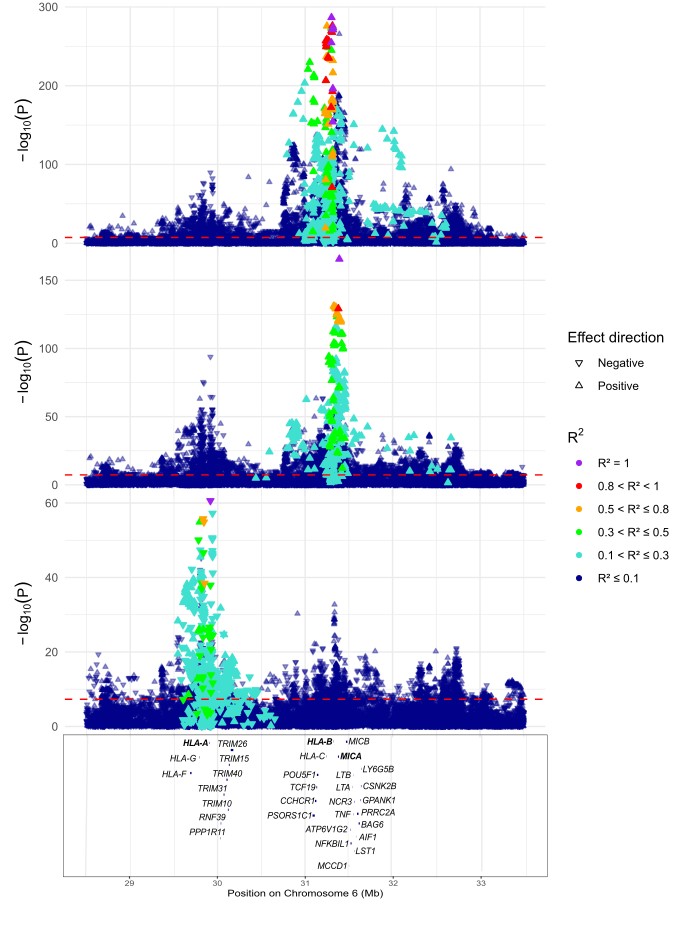

III**.**

II.

I.

**Supplementary Fig. 1b.** Locus zoom plots of the MHC region: (I) unconditioned; (II) conditioned on rs6908205; (III) conditioned on rs6908205 and rs4418214. Closest genes to lead SNPs are highlighted in bold. Triangle indicate effect direction beta (aligned to 1000 Genomes reference); color scale reflects linkage disequilibrium (R²).

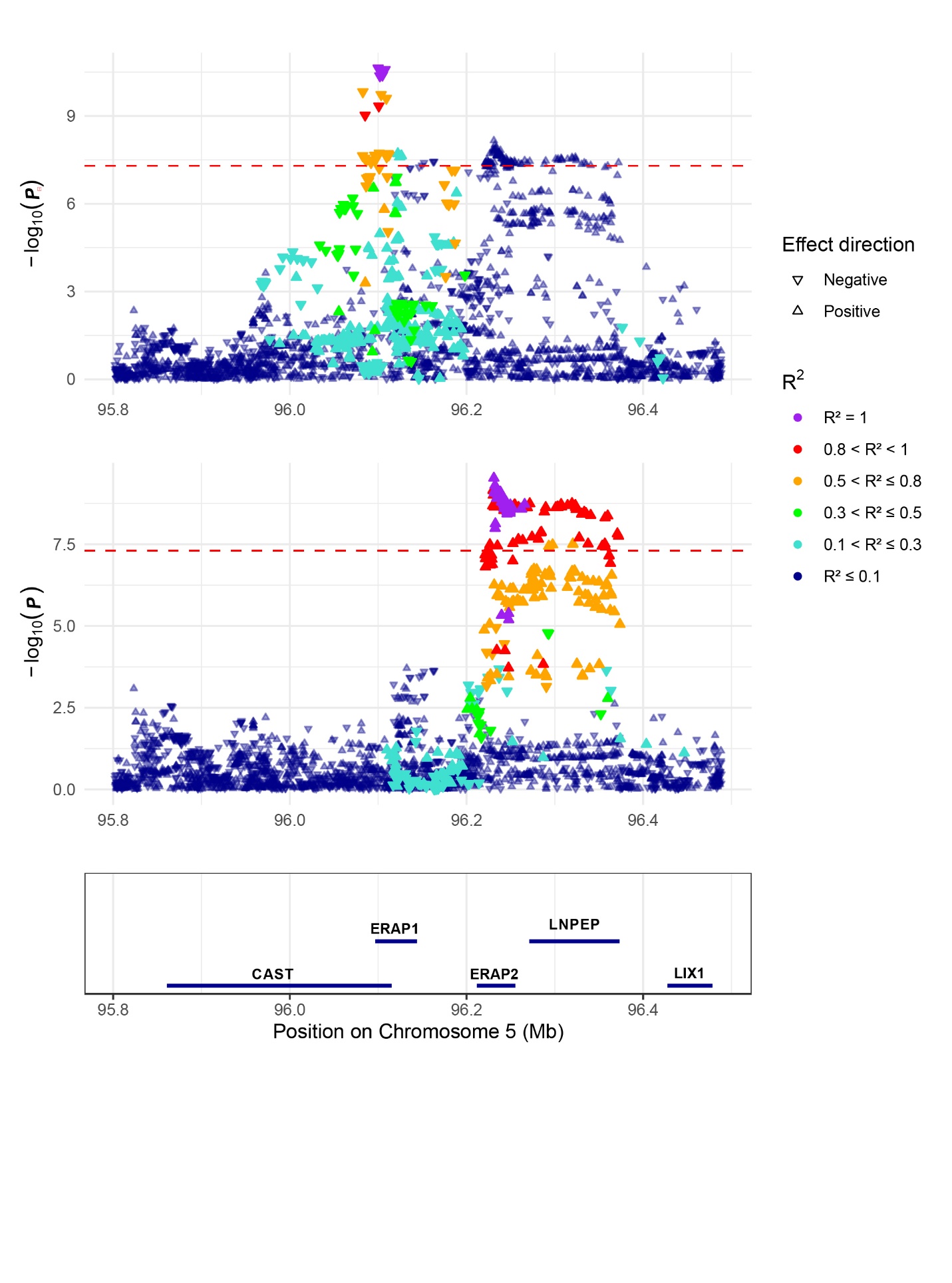

I.

II.

**Supplementary Fig. 1c.** Locus zoom plots of the *5q15* region: (I) unconditioned; (II) conditioned on rs26502. Triangle indicate effect direction beta (aligned to 1000 Genomes reference); color scale reflects linkage disequilibrium (R²).

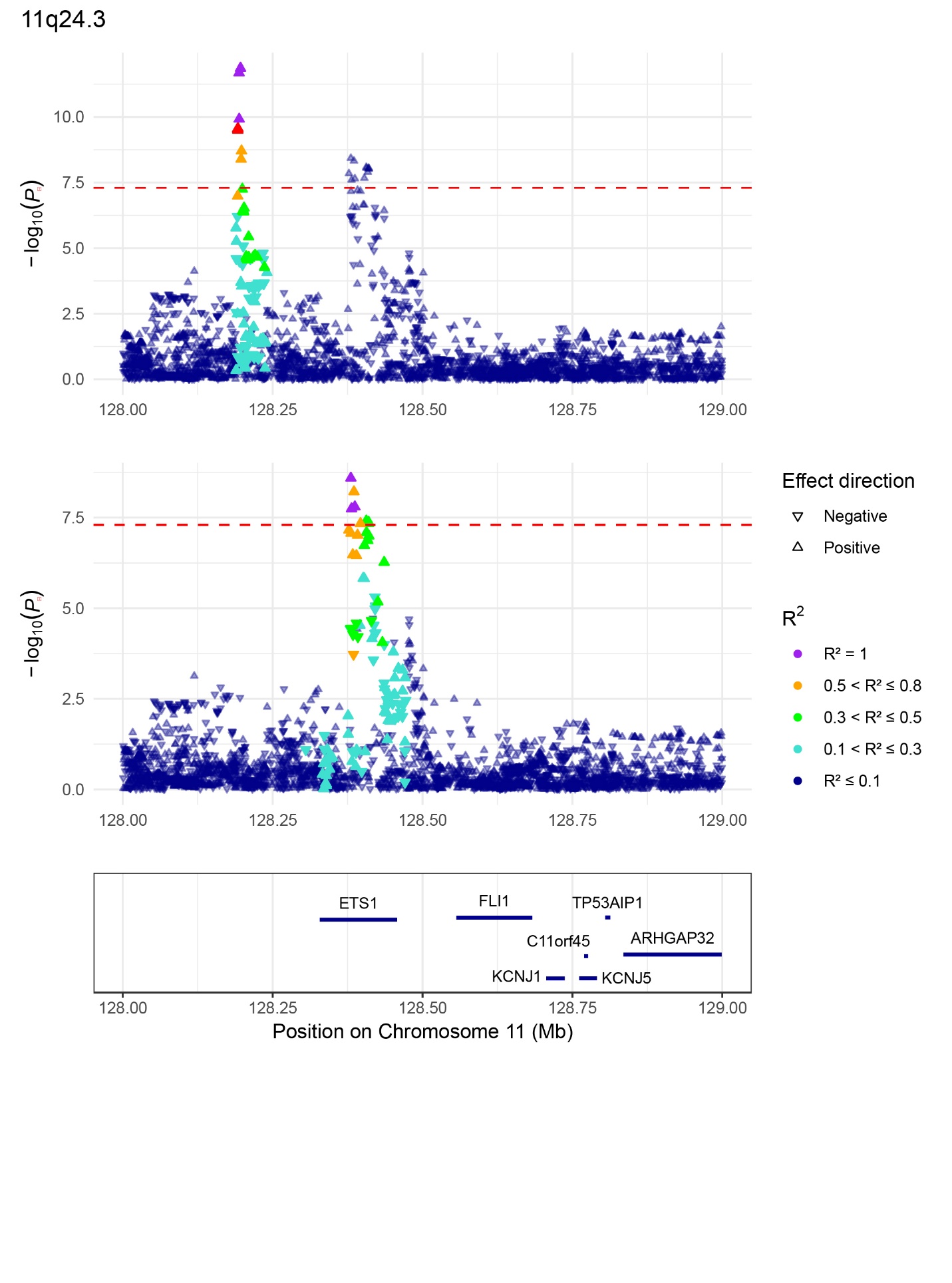

II.

I.

**Supplementary Fig. 1d.** Locus zoom plots of the *11q24.3* regions: (I) unconditioned; (II) conditioned on rs11221249. Triangle indicate effect direction beta (aligned to 1000 Genomes reference); color scale reflects linkage disequilibrium (R²).

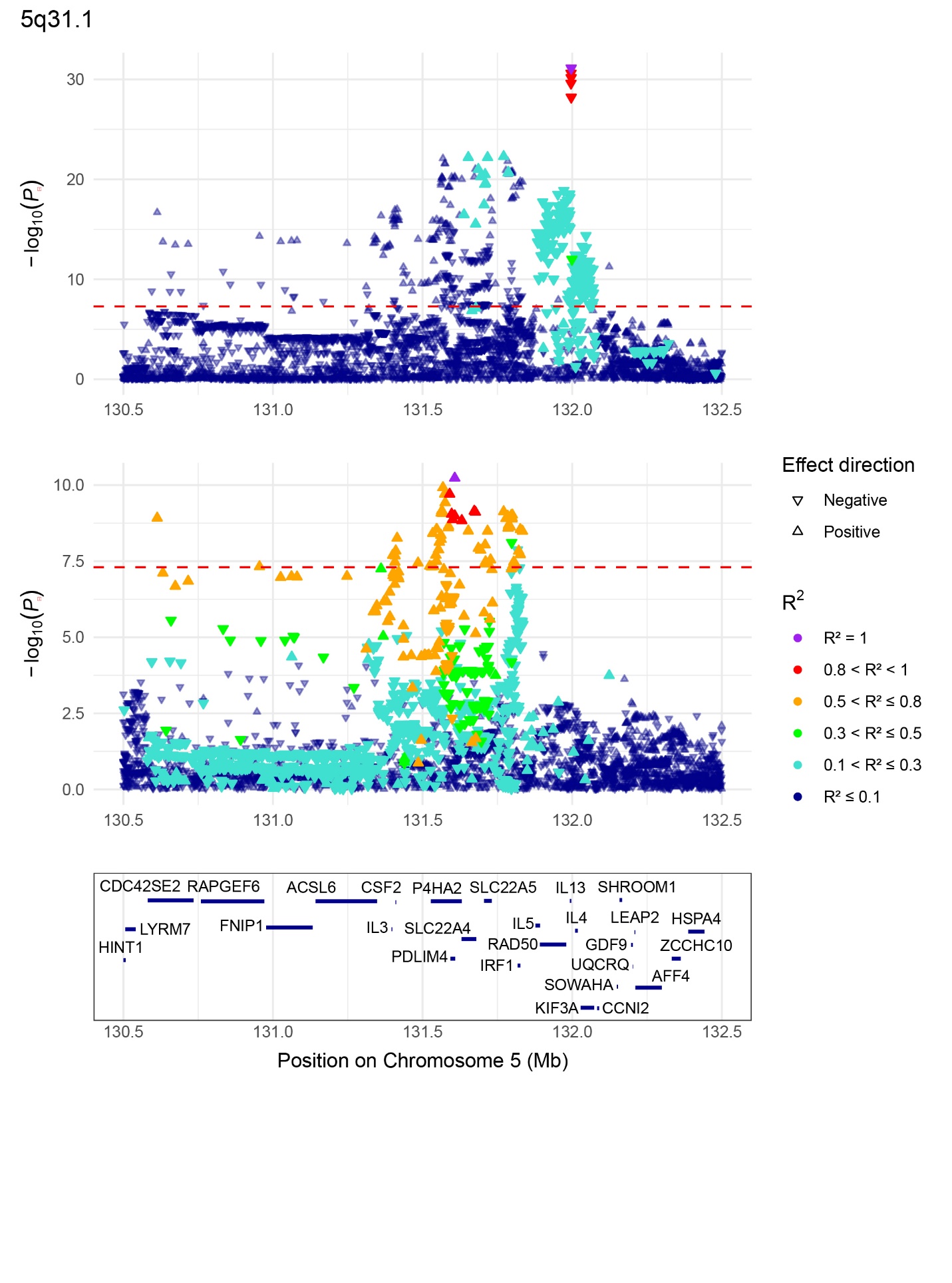

I.

II.

**Supplementary Fig. 1e.** Locus zoom plots of the *5q31.1* region: (I) unconditioned; (II) conditioned on rs2188962. Triangle indicate effect direction beta (aligned to 1000 Genomes reference); color scale reflects linkage disequilibrium (R²).

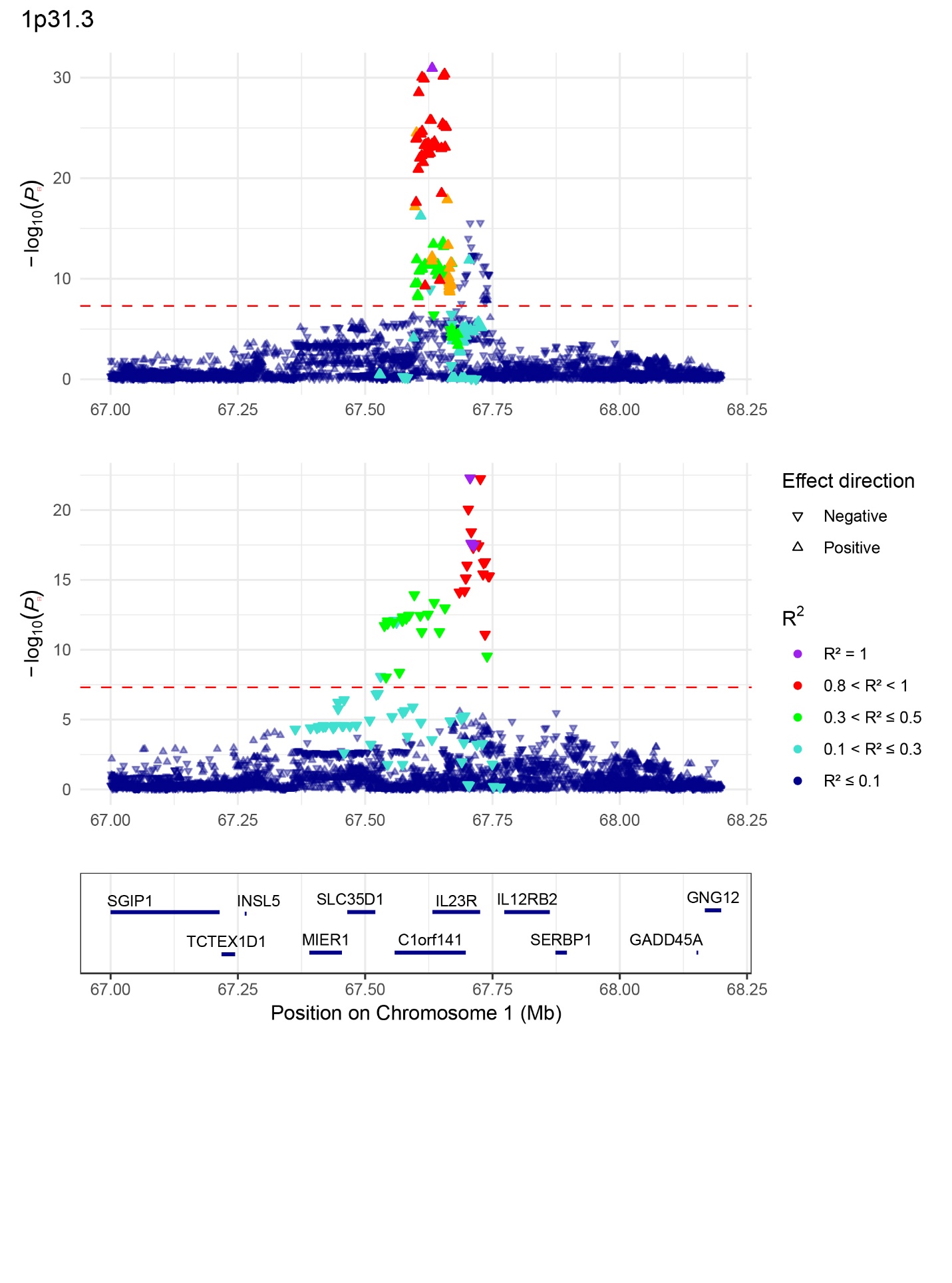

II.

I.

I.

**Supplementary Fig. 1f.** Locus zoom plots of the *1p31.3* region: (I) unconditioned; (II) conditioned on rs11465754. Triangle indicate effect direction beta (aligned to 1000 Genomes reference); color scale reflects linkage disequilibrium (R²).

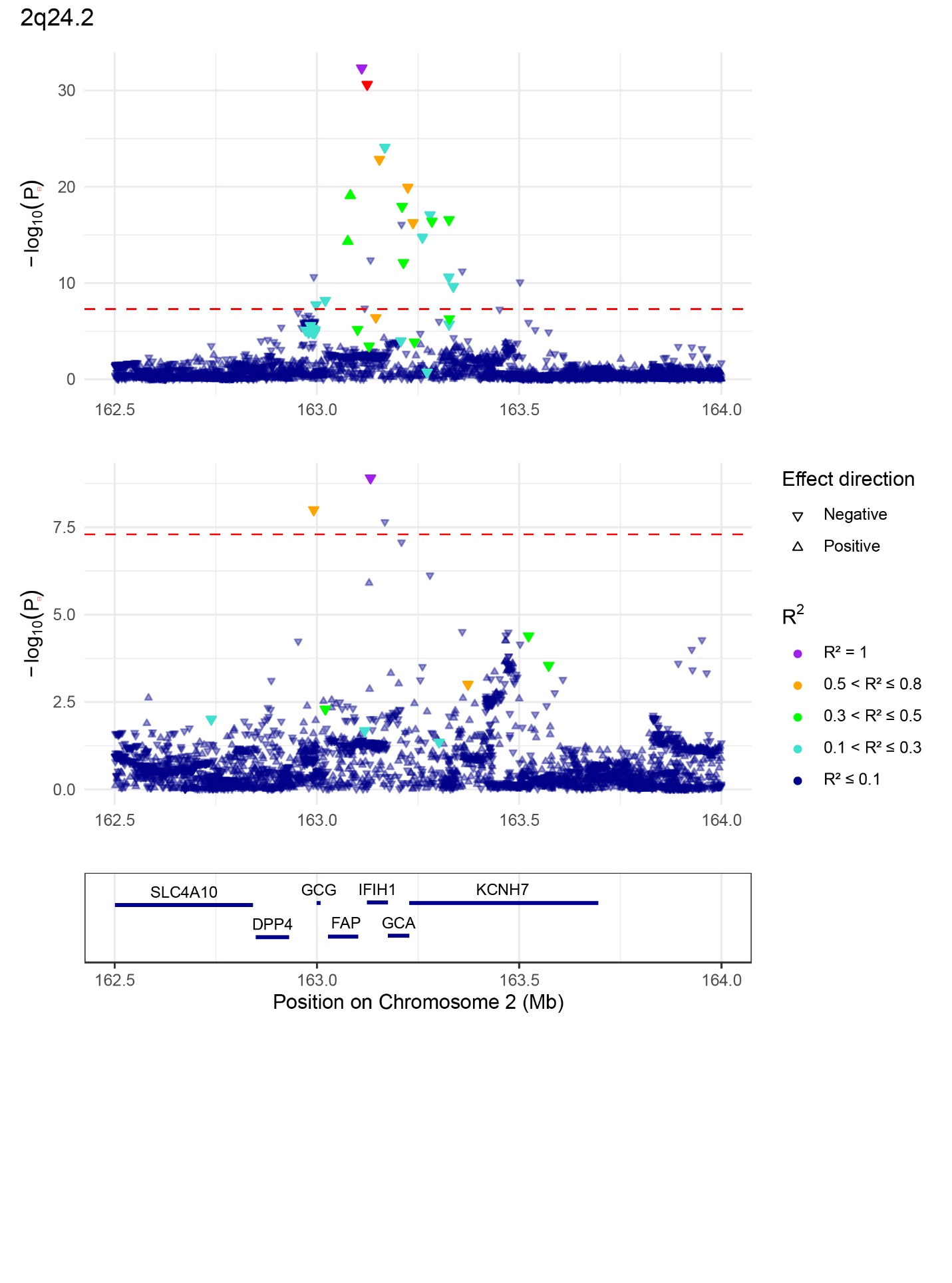

I.

II.

**Supplementary Fig. 1g.** Locus zoom plots of the *2q24.2* region: (I) unconditioned; (II) conditioned on rs2111485. Triangle indicate effect direction beta (aligned to 1000 Genomes reference); color scale reflects linkage disequilibrium (R²).

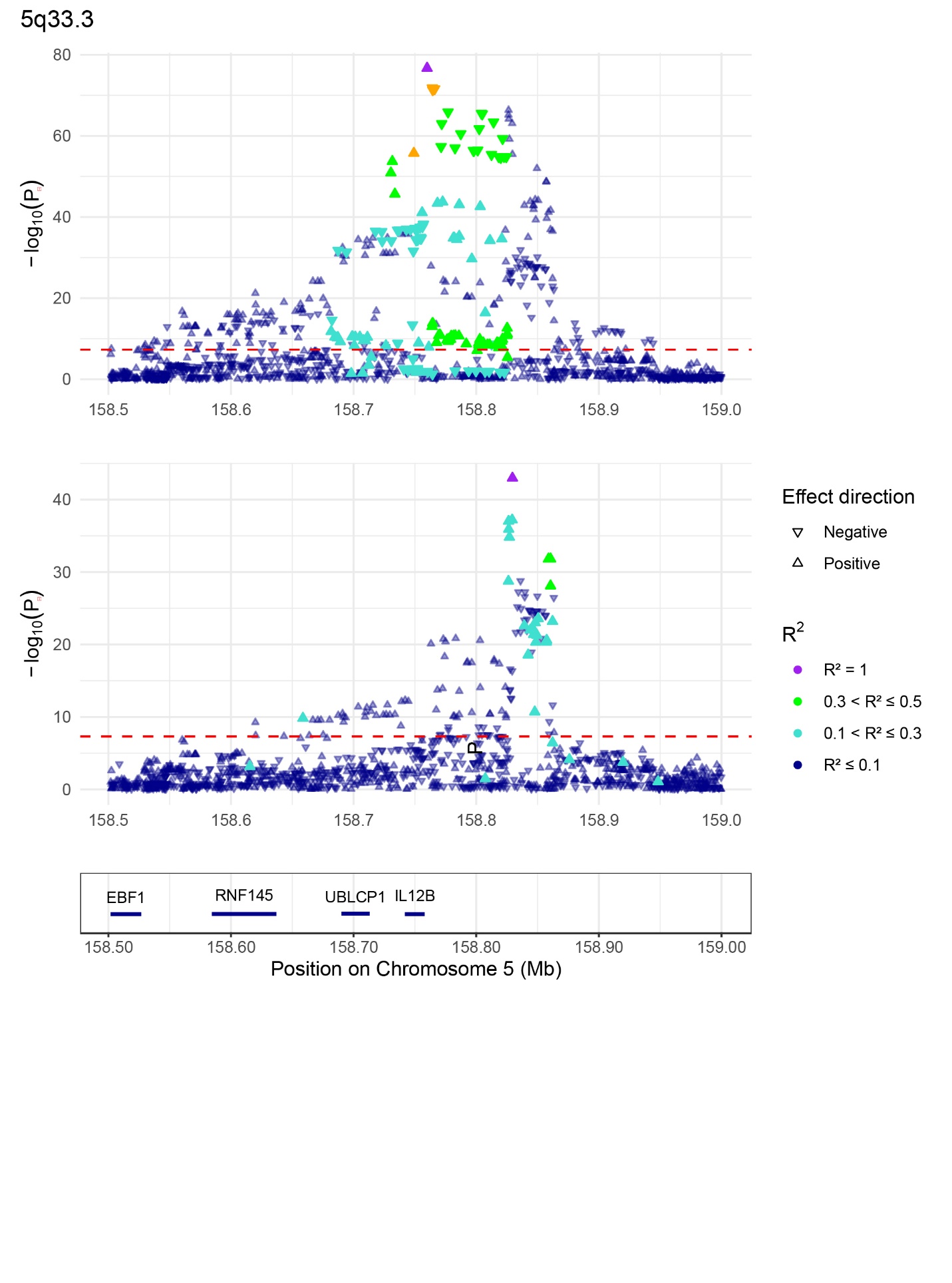

II.

I.

**Supplementary Fig. 1h.** Locus zoom plots of the *5q33.3* region: (I) unconditioned; (II) conditioned on rs2546890. Triangle indicate effect direction beta (aligned to 1000 Genomes reference); color scale reflects linkage disequilibrium (R²).

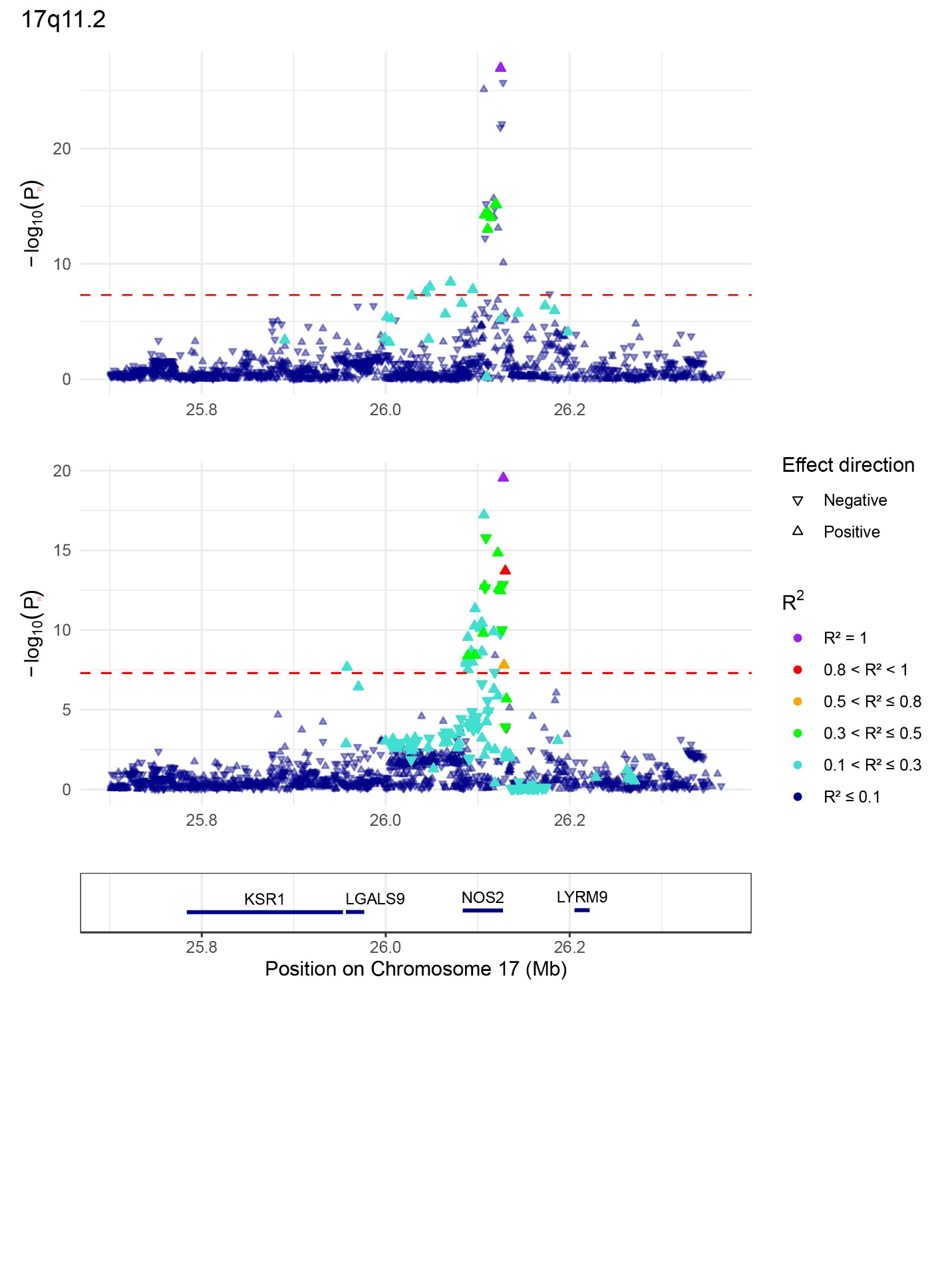
 **Supplementary Fig. 1i.** Locus zoom plots of the *17q11.2* region: (I) unconditioned; (II) conditioned on rs28998802. Triangle indicate effect direction beta (aligned to 1000 Genomes reference); color scale reflects linkage disequilibrium (R²).

II.

I.

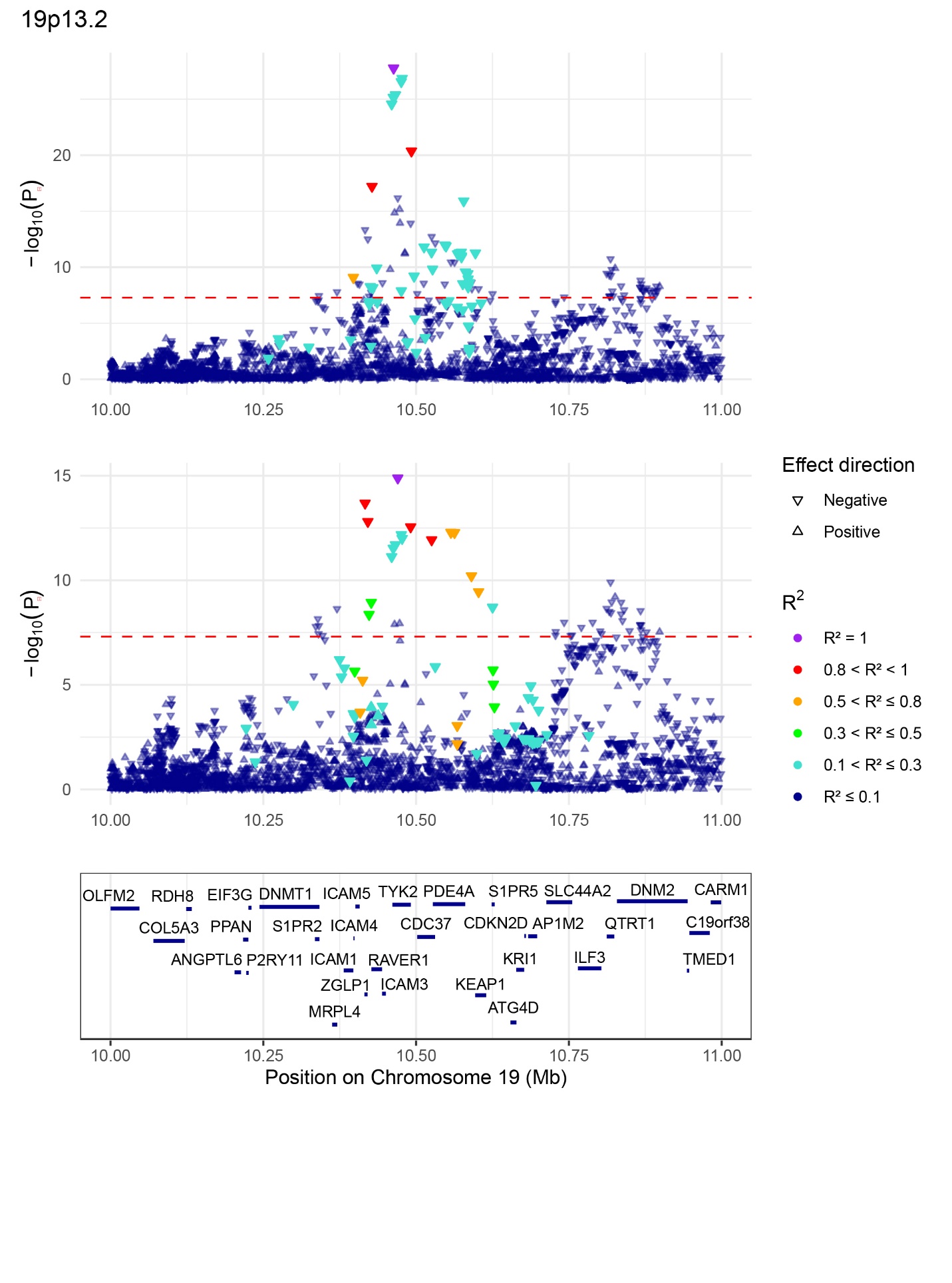

I.

II.

**Supplementary Fig. 1j.** Locus zoom plots of the *19p13.2* region: (I) unconditioned; (II) conditioned on rs34536443. Triangle indicate effect direction beta (aligned to 1000 Genomes reference); color scale reflects linkage disequilibrium (R²).

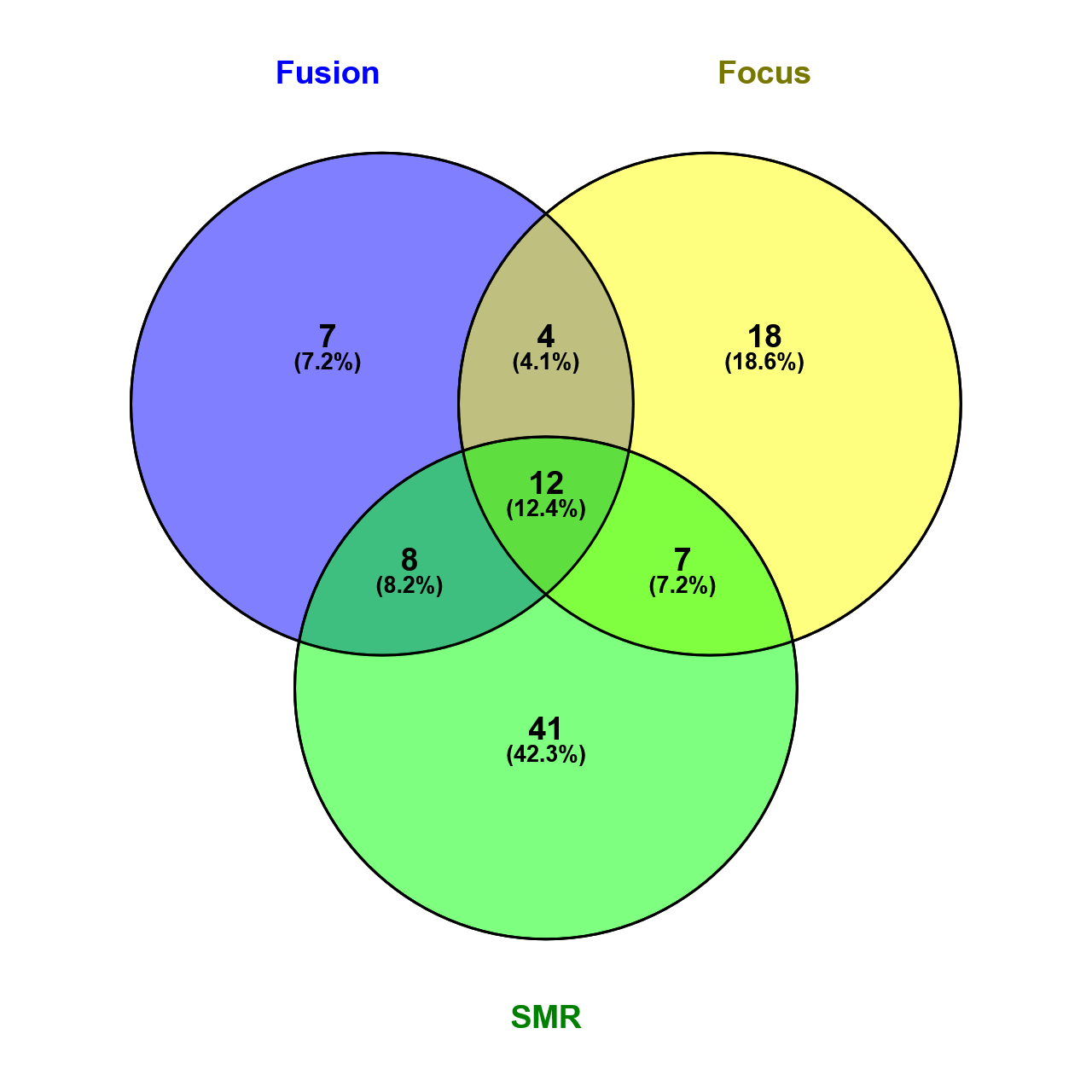

*IFNLR1, DENND1B, ERAP1, DDX58, CDHR5, RNASEH2, AP5B1, RMI2, TYK2, SPATA2, UBE2L3, CCDC116*

*B3GNT2, ERAP2, SLC22A5, RNF145, MFSD4B, PDE8A, SLC9A8*

*CDC42SE2, DRD4, PRDX5, CCDC88B, COQ10A, CNPY2, IL23A, STAT2*

*HSPA4, PHRF1, PPP1R14B, KAT5*

**Supplementary Fig. 2.** Overlap of genes prioritized by FUSION, FOCUS, and SMR. Venn diagram illustrating the overlap of genes prioritized at PsA-associated loci using three colocalization-based approaches: FUSION (blue), FOCUS (yellow), and SMR (green). Overlapping regions represent genes jointly prioritized by two or more methods, providing stronger evidence for their involvement. Numbers within each segment indicate the count of overlapping genes, while percentages reflect the proportion of shared genes between the respective methods. The names of overlapping genes are displayed in adjacent boxes for clarity.

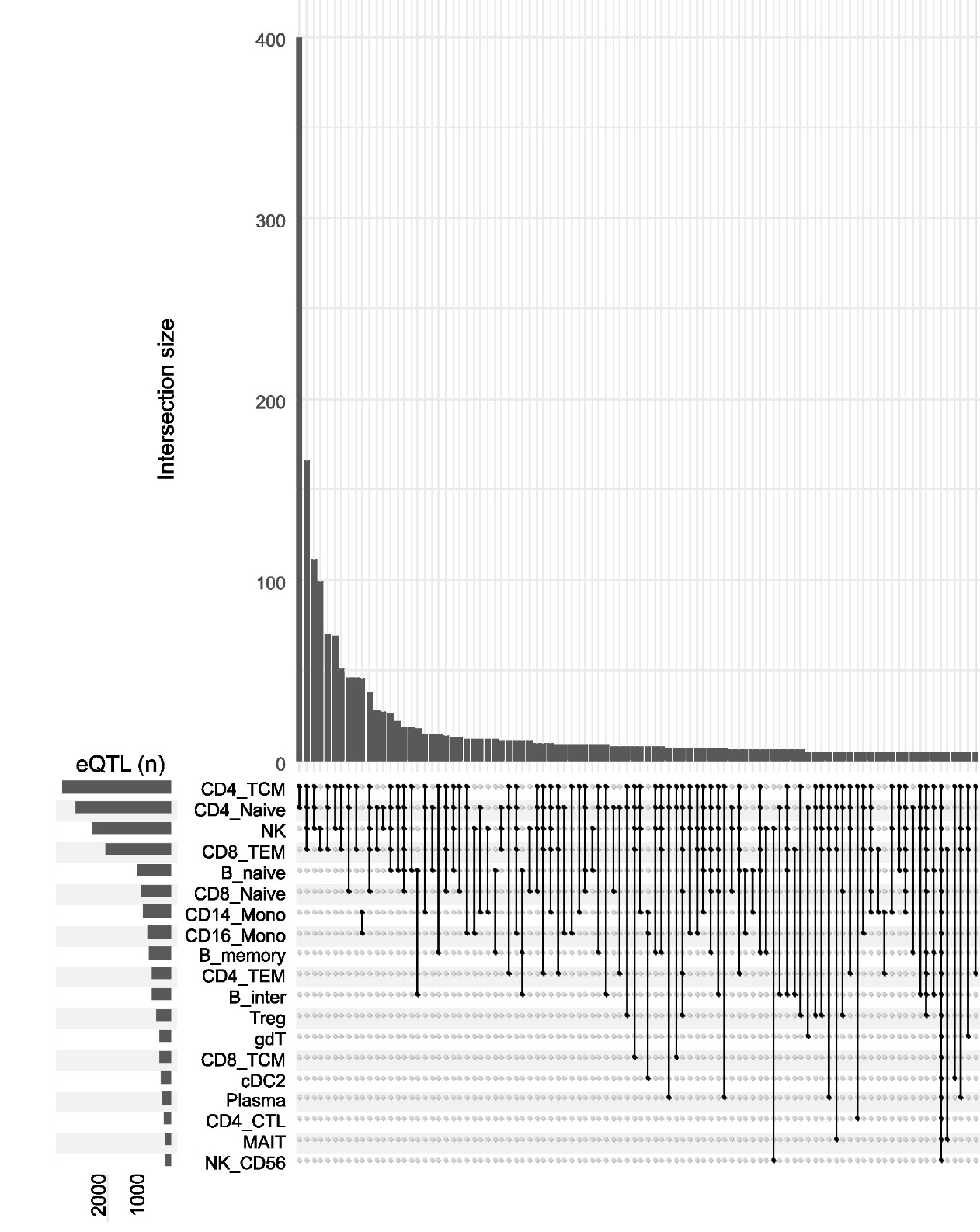

**Supplementary Figure 3. UpSet plot illustrating overlap of genes prioritized by single-cell eQTL analysis in the OneK1K cohort.** This UpSet plot displays the intersection of significant eGenes (P<1×10⁻⁵) identified across various immune cell populations in the OneK1K cohort. Horizontal bars on the left represent the total number of significant eGenes detected in each individual cell population. The matrix layout beneath the vertical bars indicates the specific cell populations involved in each intersection, with filled circles denoting inclusion. Vertical bars quantify the number of eGenes shared among the corresponding cell populations.
